## Supporting Information for "Effects of an urban sanitation intervention on childhood enteric infection and diarrhea in Maputo, Mozambique: a controlled before-and-after trial"

**Supplemental information for Effects of an urban sanitation intervention on childhood enteric infection and diarrhea in Maputo, Mozambique**

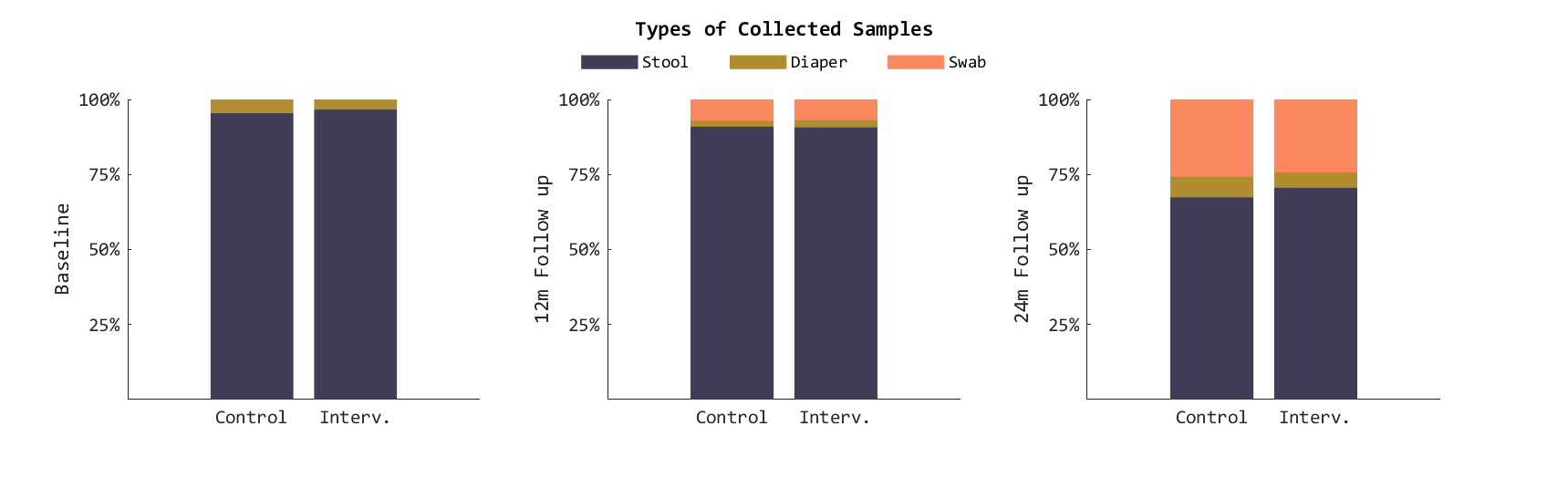

Supplemental Figure 1: Proportion of each type of sample collected during the baseline, 12-month, and 24-month phases. Results stratified by study arm. Rectal swabs were not introduced until the 12-month phase of the study. Source files available in Supplemental Figure 1 - source data 1 and Supplemental Figure 1 - source code 1.

### Supplemental Table 1: Number and proportion of sample types collected in each arm at each phase.

|  | Baseline | | 12-month | | 24-month | |
| --- | --- | --- | --- | --- | --- | --- |
|  | Control | Intervention | Control | Intervention | Control | Intervention |
| Whole stool | 377 (96%) | 351 (97%) | 361 (91%) | 380 (93%) | 307 (67%) | 333 (72%) |
| Diarrheal diaper | 15 (3.8%) | 10 (2.8%) | 4 (1.0%) | 4 (0.98%) | 32 (7.0%) | 20 (4.3%) |
| Rectal swab* | 0 (0%) | 0 (0%) | 30 (7.6%) | 24 (5.9%) | 120 (26%) | 109 (24%) |

* Mean concentration of double-stranded DNA recovered from whole stool was 40.8 ng/μL (SD=16.5, n=33 with 57 samples excluded as their concentrations exceeded the upper detection limit of the assay), diaper samples was 28.7 ng/μL (SD=16.9, n=61 with 16 samples excluded as concentrations exceeded upper detection limit of assay), and from rectal swabs was 26.3 ng/μL (SD=15.5, n=195 with 25 samples excluded as concentrations exceeded upper detection limit of assay). Only a subset of each sample type assayed for dsDNA concentration. Source files available in Supplemental Table 1 - source data 1 and Supplemental Table 1 - source code 1.

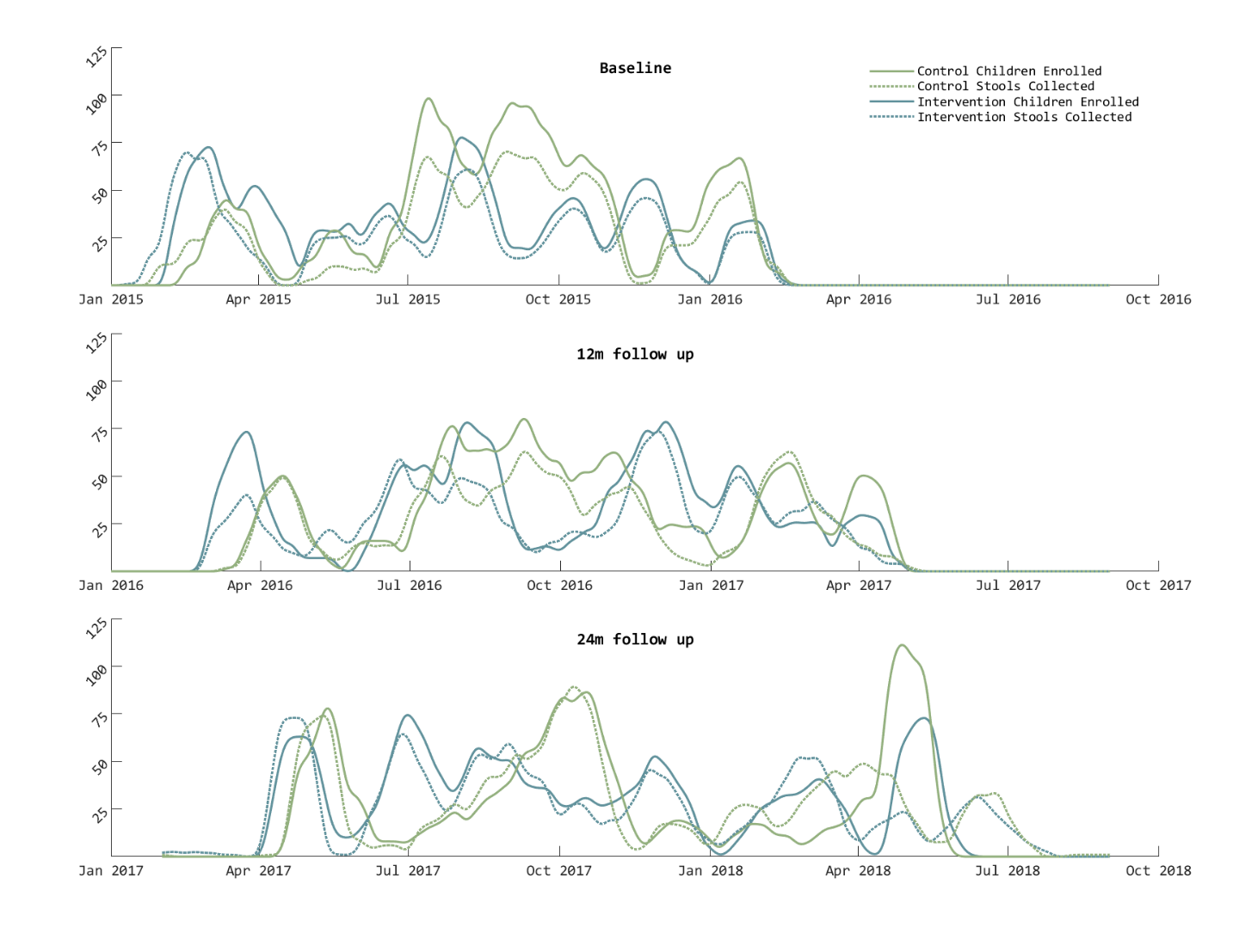

Supplemental Figure 2: Enrollment and stool sample collection profile. Graphs depict four week rolling average of the number of intervention and control children enrolled/visited (solid lines) and the number of stool samples collected (including whole stool, diaper samples, and rectal swabs) during the baseline, 12-month, and 24-month phases. The overall success of stool sample collection was 78% at baseline, 86% at 12-month, and 90% at 24-month. The increase in success rate was due to the introduction of rectal swab collection during the 12-month phase. Source files available in Supplemental Figure 2 - source data 1 and Supplemental Figure 2 - source code 1.

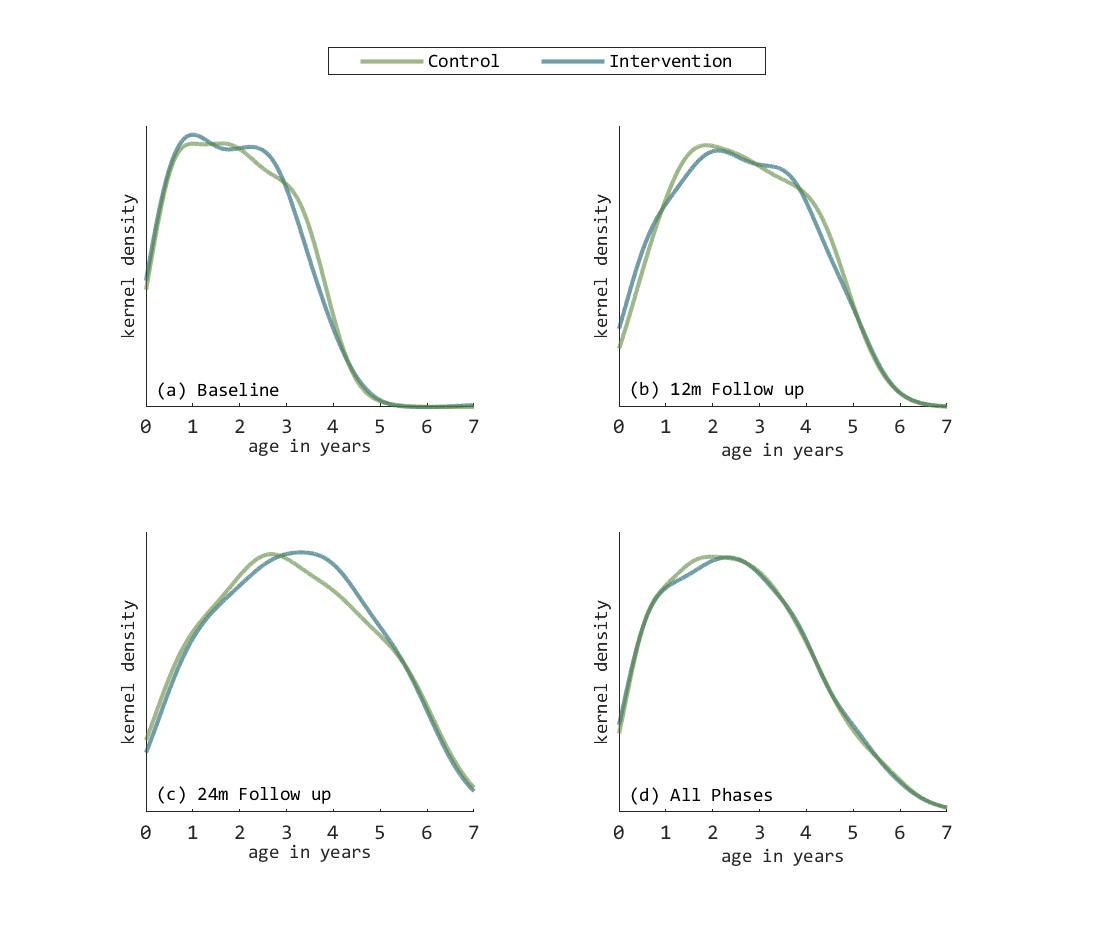

Supplemental Figure 3: Distribution of age (years) of enrolled children at each phase. Results are presented as kernel density plots and stratified by study arm (intervention=blue, control=green) and phase: (a) Baseline phase, (b) 12-month follow-up, (c) 24-month follow-up, and (d) All phases combined. Source files available in Supplemental Figure 3 - source data 1 and Supplemental Figure 3 - source code 1.

### Supplemental Table 2: Age stratified baseline prevalence of health outcomes.

|  | **Baseline Prevalence** | | |
| --- | --- | --- | --- |
|  | **1 - 11 months** | **12-23 months** | **24 - 48 months** |
| Any bacterial or protozoan infection |  |  |  |
| All children | 108/208 (52%) | 179/221 (81%) | 277/297 (93%) |
| Control | 57/109 (52%) | 101/119 (85%) | 143/152 (94%) |
| Intervention | 51/99 (52%) | 78/102 (76%) | 134/145 (92%) |
| Any STH infection |  |  |  |
| All children | 30/185 (16%) | 89/203 (44%) | 171/277 (62%) |
| Control | 17/93 (18%) | 50/112 (45%) | 94/144 (65%) |
| Intervention | 13/92 (14%) | 39/91 (43%) | 77/133 (58%) |
| Diarrhea |  |  |  |
| All children | 37/258 (14%) | 52/264 (20%) | 36/427 (8.4%) |
| Control | 19/138 (14%) | 27/146 (18%) | 20/234 (8.6%) |
| Intervention | 18/120 (15%) | 25/118 (21%) | 16/193 (8.3%) |
| Any bacterial infection |  |  |  |
| All children | 94/208 (45%) | 150/221 (68%) | 229/297 (77%) |
| Intervention | 53/109 (49%) | 89/119 (75%) | 117/152 (77%) |
| All children | 41/99 (41%) | 61/102 (60%) | 112/145 (77%) |
| *Shigella* |  |  |  |
| All children | 19/208 (9.1%) | 97/221 (44%) | 192/297 (65%) |
| Control | 10/109 (9.2%) | 57/119 (48%) | 101/152 (66%) |
| Intervention | 9/99 (9.1%) | 40/102 (39%) | 91/145 (63%) |
| ETEC |  |  |  |
| All children | 47/208 (23%) | 81/221 (37%) | 90/297 (30%) |
| Control | 25/109 (23%) | 45/119 (38%) | 43/152 (28%) |
| Intervention | 22/99 (22%) | 36/102 (35%) | 47/145 (32%) |
| *Campylobacter* |  |  |  |
| All children | 22/208 (11%) | 19/221 (8.6%) | 16/297 (5.4%) |
| Control | 14/109 (13%) | 13/119 (11%) | 10/152 (6.6%) |
| Intervention | 8/99 (8.1%) | 6/102 (5.9%) | 6/145 (4.1%) |
| *C. difficile* |  |  |  |
| All children | 23/208 (11%) | 10/221 (4.5%) | 2/297 (0.67%) |
| Control | 13/109 (12%) | 7/119 (5.9%) | 2/152 (1.3%) |
| Intervention | 10/99 (10%) | 3/102 (2.9%) | 0/145 (0.0%) |
| *E. coli* o157 |  |  |  |
| All children | 6/208 (2.9%) | 10/221 (4.5%) | 15/297 (5%) |
| Control | 4/109 (3.7%) | 3/119 (2.5%) | 6/152 (4%) |
| Intervention | 2/99 (2%) | 7/102 (6.9%) | 9/145 (6.2%) |
| STEC |  |  |  |
| All children | 3/208 (1.4%) | 7/221 (3.2%) | 3/297 (1%) |
| Control | 0/109 (0.0%) | 1/119 (0.84%) | 2/152 (1.3%) |
| Intervention | 3/99 (3%) | 6/102 (5.9%) | 1/145 (0.69%) |
| *Y.* *enterocolitica* |  |  |  |
| All children | 0/208 (0.0%) | 1/221 (0.45%) | 0/297 (0.0%) |
| Control | 0/109 (0.0%) | 0/119 (0.0%) | 0/152 (0.0%) |
| Intervention | 0/99 (0.0%) | 1/102 (0.98%) | 0/145 (0.0%) |
| *V. cholerae* |  |  |  |
| All children | 0/208 (0.0%) | 0/221 (0.0%) | 0/297 (0.0%) |
| Control | 0/109 (0.0%) | 0/119 (0.0%) | 0/152 (0.0%) |
| Intervention | 0/99 (0.0%) | 0/102 (0.0%) | 0/145 (0.0%) |
| Any Protozoa |  |  |  |
| All children | 36/208 (17%) | 120/221 (54%) | 223/297 (75%) |
| Control | 14/109 (13%) | 68/119 (57%) | 114/152 (75%) |
| Intervention | 22/99 (22%) | 52/102 (51%) | 109/145 (75%) |
| *Giardia* |  |  |  |
| All children | 28/208 (13%) | 119/221 (54%) | 219/297 (74%) |
| Control | 12/109 (11%) | 67/119 (56%) | 113/152 (74%) |
| Intervention | 16/99 (16%) | 52/102 (51%) | 106/145 (73%) |
| *Cryptosporidium* |  |  |  |
| All children | 10/208 (4.8%) | 9/221 (4.1%) | 5/297 (1.7%) |
| Control | 2/109 (1.8%) | 5/119 (4.2%) | 1/152 (0.66%) |
| Intervention | 8/99 (8.1%) | 4/102 (3.9%) | 4/145 (2.8%) |
| *E. histolytica* |  |  |  |
| All children | 1/208 (0.48%) | 0/221 (0.0%) | 3/297 (1%) |
| Control | 0/109 (0.0%) | 0/119 (0.0%) | 0/152 (0.0%) |
| Intervention | 1/99 (1%) | 0/102 (0.0%) | 3/145 (2.1%) |
| Any virus |  |  |  |
| All children | 36/208 (17%) | 34/221 (15%) | 33/297 (11%) |
| Control | 15/109 (14%) | 19/119 (16%) | 19/152 (13%) |
| Intervention | 21/99 (21%) | 15/102 (15%) | 14/145 (9.7%) |
| Norovirus GI/GII |  |  |  |
| All children | 27/208 (13%) | 25/221 (11%) | 23/297 (7.7%) |
| Control | 12/109 (11%) | 14/119 (12%) | 12/152 (7.9%) |
| Intervention | 15/99 (15%) | 11/102 (11%) | 11/145 (7.6%) |
| Adenovirus 40/41 |  |  |  |
| All children | 7/208 (3.4%) | 7/221 (3.2%) | 8/297 (2.7%) |
| Control | 4/109 (3.7%) | 3/119 (2.5%) | 6/152 (4%) |
| Intervention | 3/99 (3%) | 4/102 (3.9%) | 2/145 (1.4%) |
| Rotavirus A |  |  |  |
| All children | 3/208 (1.4%) | 5/221 (2.3%) | 2/297 (0.67%) |
| Control | 0/109 (0.0%) | 2/119 (1.7%) | 1/152 (0.66%) |
| Intervention | 3/99 (3%) | 3/102 (2.9%) | 1/145 (0.69%) |
| Coinfection, ≥2 GPP pathogens |  |  |  |
| All children | 48/208 (23%) | 118/221 (53%) | 203/297 (68%) |
| Control | 23/109 (21%) | 69/119 (58%) | 104/152 (68%) |
| Intervention | 25/99 (25%) | 49/102 (48%) | 99/145 (68%) |
| *Trichuris* |  |  |  |
| All children | 20/185 (11%) | 69/203 (34%) | 150/277 (54%) |
| Control | 10/93 (11%) | 38/112 (34%) | 82/144 (57%) |
| Intervention | 10/92 (11%) | 31/91 (34%) | 68/133 (51%) |
| *Ascaris* |  |  |  |
| All children | 21/185 (11%) | 53/203 (26%) | 81/277 (29%) |
| Control | 12/93 (13%) | 33/112 (29%) | 47/144 (33%) |
| Intervention | 9/92 (9.8%) | 20/91 (22%) | 34/133 (26%) |
| Coinfection, ≥2 STH |  |  |  |
| All children | 11/185 (6%) | 33/203 (16%) | 60/277 (22%) |
| Control | 5/93 (5.4%) | 21/112 (19%) | 35/144 (24%) |
| Intervention | 6/92 (6.5%) | 12/91 (13%) | 25/133 (19%) |
| Number of GPP infections |  |  |  |
| All children | 0.94 (1.1) | 1.8 (1.2) | 1.9 (0.95) |
| Control | 0.88 (1.1) | 1.8 (1.1) | 2 (0.93) |
| Intervention | 1 (1.1) | 1.7 (1.3) | 1.9 (0.98) |
| Number of STH infections |  |  |  |
| All children | 0.23 (0.55) | 0.61 (0.75) | 0.86 (0.76) |
| Control | 0.24 (0.54) | 0.64 (0.78) | 0.9 (0.76) |
| Intervention | 0.23 (0.56) | 0.57 (0.72) | 0.8 (0.76) |

Data presented n/N (%) or mean (standard deviation). All bacterial, protozoan, and viral pathogens were measured using the Luminex Gastrointestinal Pathogen panel. STH were measured using the Kato-Katz method. Diarrhea was measured via caregiver report in household surveys. Source files available in Supplemental Table 2 - source data 1 and Supplemental Table 2 - source code 1.

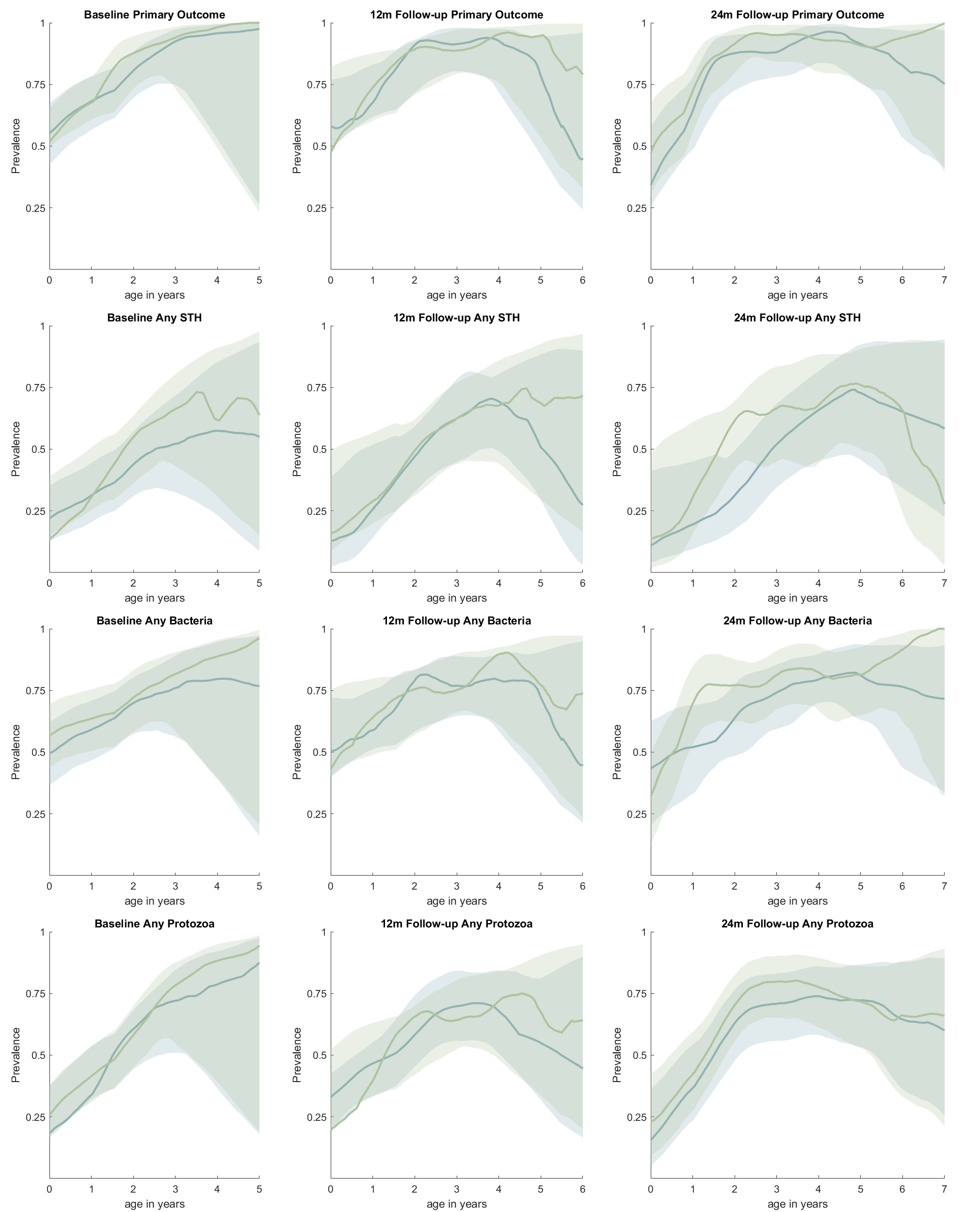

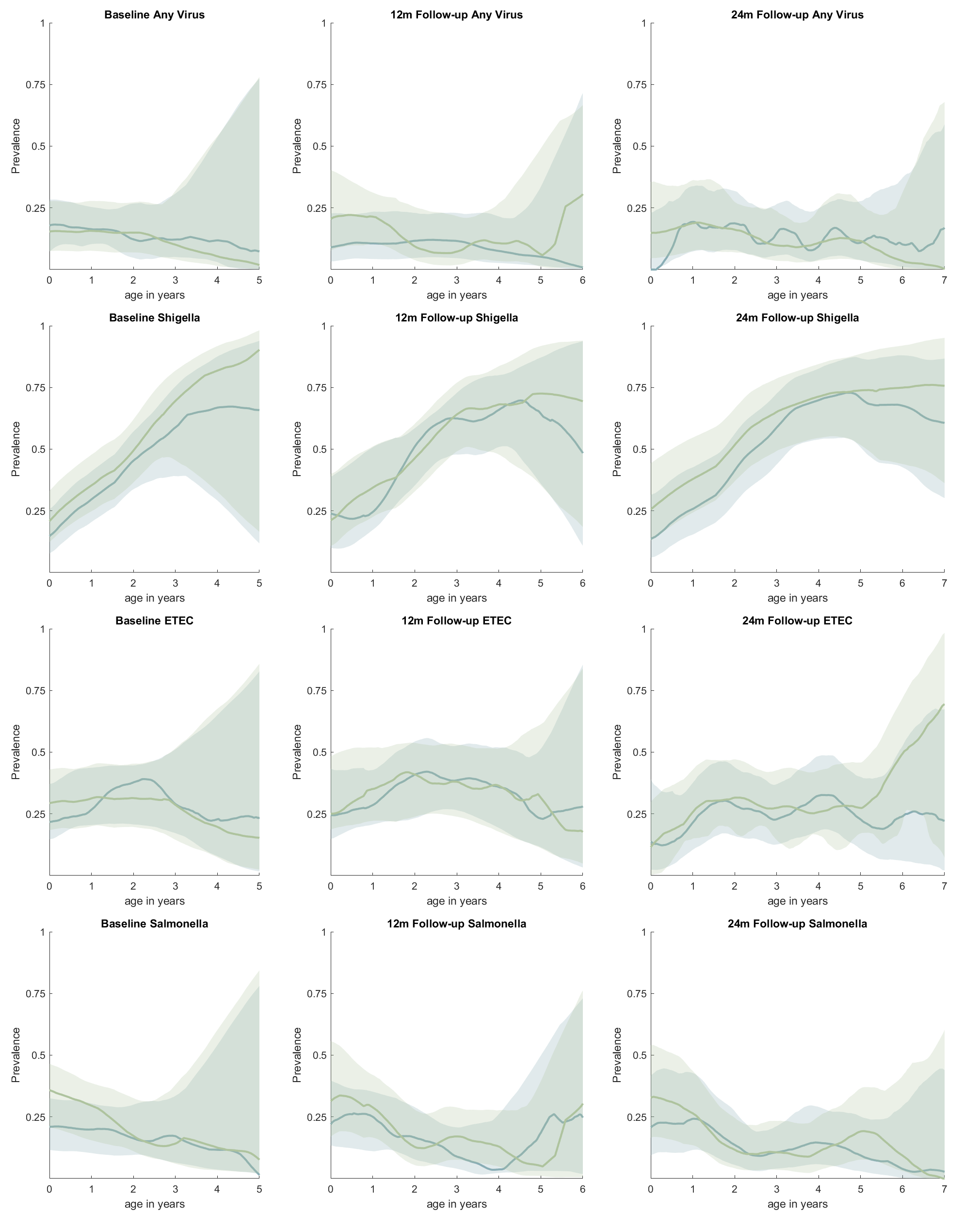

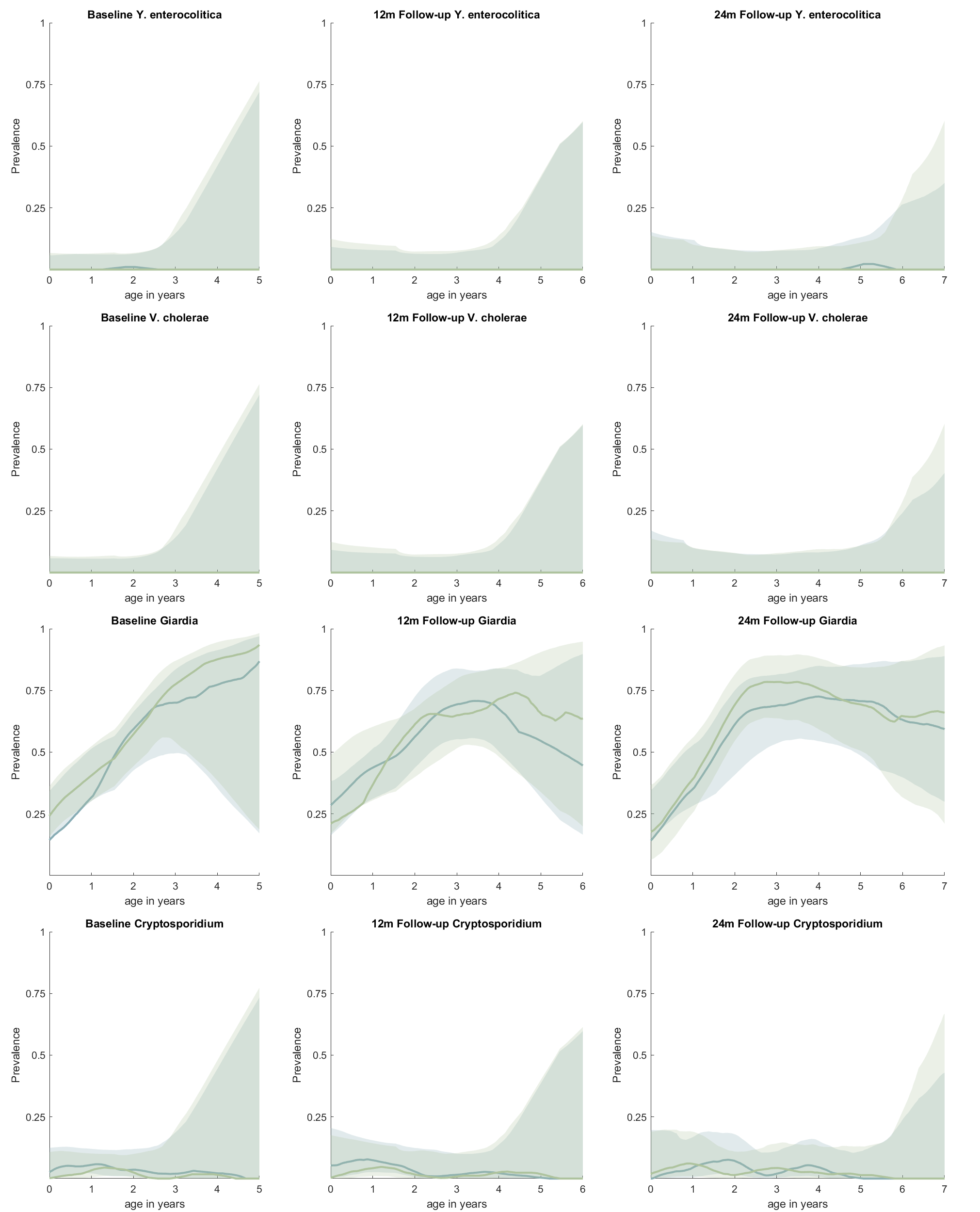

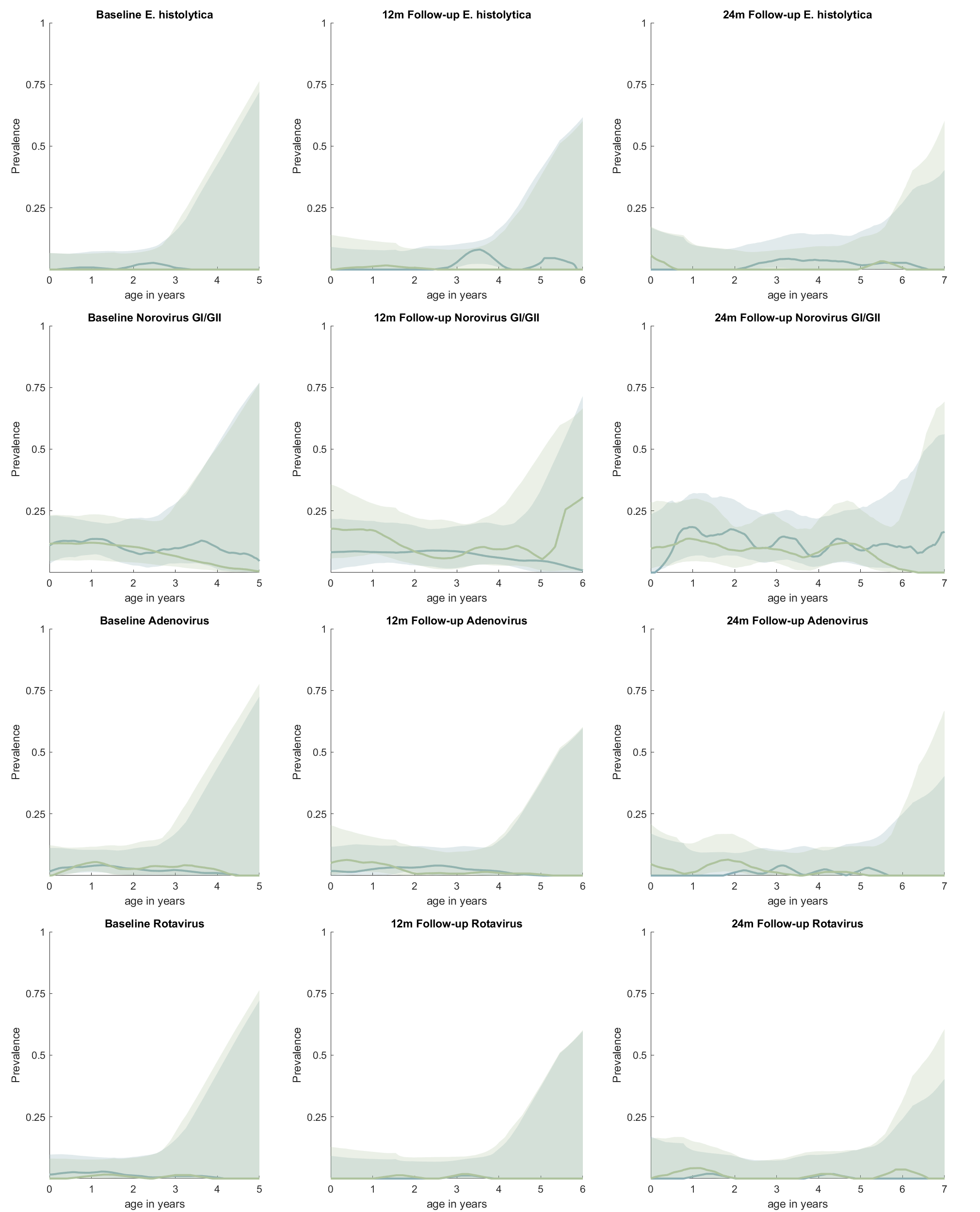

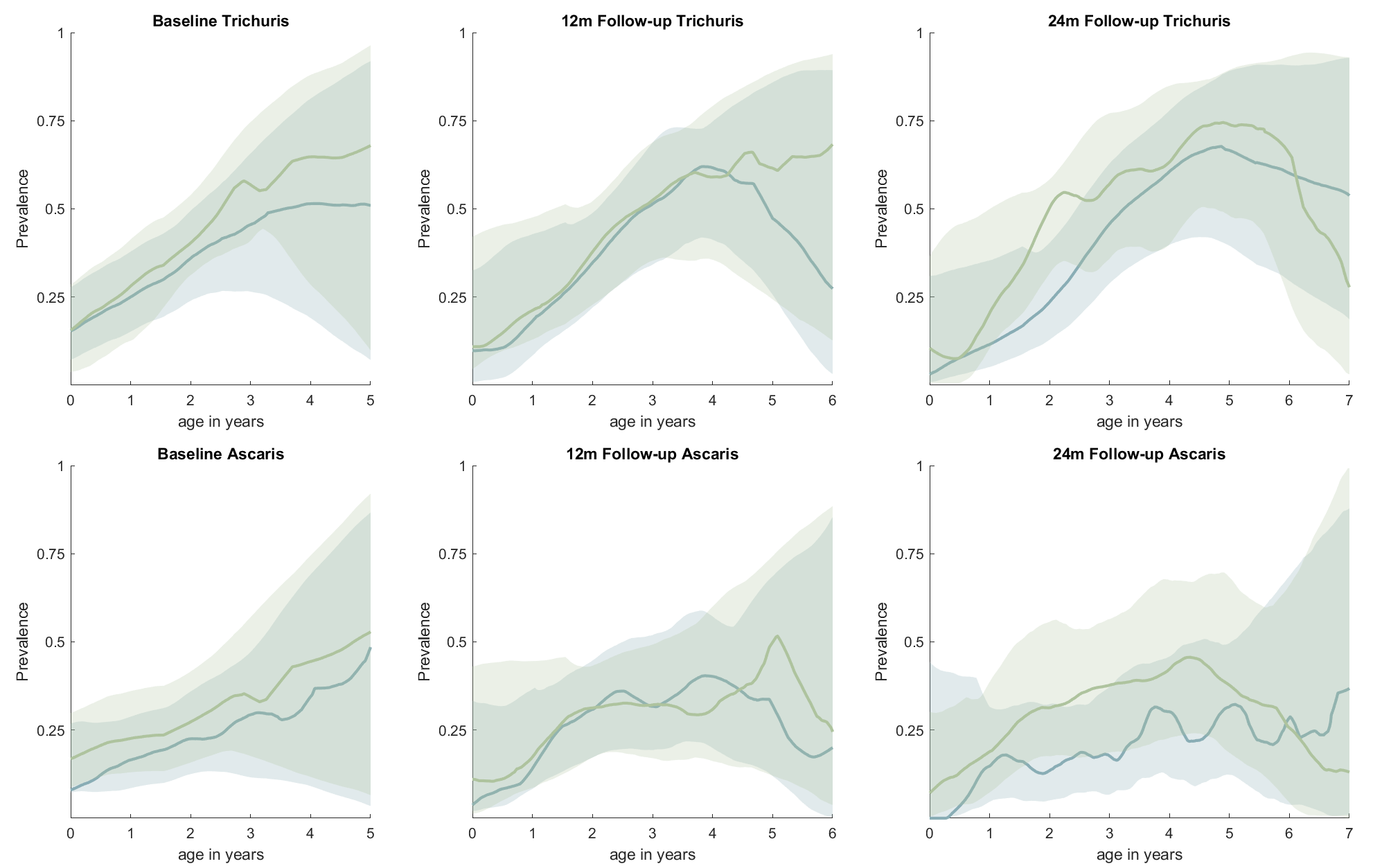

Supplemental Figure 4: Prevalence of pathogens by age at baseline, 12-month, and 24-month phases. Results are smoothed averages stratified by study arm with 95% confidence intervals represented by shaded areas. Source files available in Supplemental Figure 4 - source data 1 and Supplemental Figure 4 - source code 1.

Supplemental Table 3: Baseline enrollment characteristics of children with and without repeated measures at the 12-month phase. Results are presented for all children combined and stratified by study arm.

|  | All children | | | Control | | | Intervention | | |
| --- | --- | --- | --- | --- | --- | --- | --- | --- | --- |
|  | BL & 12M* | BL only† | Std. Diff.‡ | BL & 12M | BL only | Std. Diff. | BL & 12M | BL only | Std. Diff. |
| **Outcomes** | | | | | | | | | |
| Diarrhea | 83/609 (14%) | 43/365 (12%) | 0.06 | 38/310 (12%) | 29/216 (13%) | 0.03 | 45/299 (15%) | 14/149 (9.4%) | 0.17 |
| Any bacterial or protozoan infection | 376/485 (78%) | 215/268 (80%) | 0.07 | 184/234 (79%) | 129/158 (82%) | 0.08 | 192/251 (76%) | 86/110 (78%) | 0.04 |
| Any GPP infection | 390/485 (80%) | 225/268 (84%) | 0.09 | 188/234 (80%) | 135/158 (85%) | 0.14 | 202/251 (80%) | 90/110 (82%) | 0.03 |
| Any bacterial infection | 311/485 (64%) | 187/268 (70%) | 0.12 | 157/234 (67%) | 114/158 (72%) | 0.11 | 154/251 (61%) | 73/110 (66%) | 0.10 |
| *Shigella* | 200/485 (41%) | 131/268 (49%) | 0.15 | 101/234 (43%) | 78/158 (49%) | 0.12 | 99/251 (39%) | 53/110 (48%) | 0.18 |
| ETEC | 147/485 (30%) | 79/268 (29%) | 0.02 | 68/234 (29%) | 48/158 (30%) | 0.03 | 79/251 (31%) | 31/110 (28%) | 0.07 |
| *Campylobacter* | 37/485 (7.6%) | 23/268 (8.6%) | 0.03 | 22/234 (9.4%) | 17/158 (11%) | 0.05 | 15/251 (6%) | 6/110 (5.5%) | 0.02 |
| *C. difficile* | 23/485 (4.7%) | 12/268 (4.5%) | 0.01 | 15/234 (6.4%) | 7/158 (4.4%) | 0.09 | 8/251 (3.2%) | 5/110 (4.5%) | 0.07 |
| *E. coli* O157 | 19/485 (3.9%) | 12/268 (4.5%) | 0.03 | 9/234 (3.9%) | 4/158 (2.5%) | 0.07 | 10/251 (4%) | 8/110 (7.3%) | 0.14 |
| STEC | 7/485 (1.4%) | 6/268 (2.2%) | 0.06 | 1/234 (0.43%) | 2/158 (1.3%) | 0.09 | 6/251 (2.4%) | 4/110 (3.6%) | 0.07 |
| Any protozoan infection | 257/485 (53%) | 143/268 (53%) | 0.01 | 126/234 (54%) | 79/158 (50%) | 0.08 | 131/251 (52%) | 64/110 (58%) | 0.12 |
| *Giardia* | 247/485 (51%) | 140/268 (52%) | 0.03 | 122/234 (52%) | 79/158 (50%) | 0.04 | 125/251 (50%) | 61/110 (55%) | 0.11 |
| *Cryptosporidium* | 20/485 (4.1%) | 4/268 (1.5%) | 0.16 | 7/234 (3%) | 1/158 (0.63%) | 0.18 | 13/251 (5.2%) | 3/110 (2.7%) | 0.13 |
| *E. histolytica* | 2/485 (0.41%) | 2/268 (0.75%) | 0.04 | 0/234 (0.0%) | 0/158 (0.0%) | ..⁑ | 2/251 (0.80%) | 2/110 (1.8%) | 0.09 |
| Any viral infection | 66/485 (14%) | 39/268 (15%) | 0.03 | 31/234 (13%) | 22/158 (14%) | 0.02 | 35/251 (14%) | 17/110 (15%) | 0.04 |
| Adenovirus 40/41 | 14/485 (2.9%) | 8/268 (3%) | 0.01 | 8/234 (3.4%) | 5/158 (3.2%) | 0.01 | 6/251 (2.4%) | 3/110 (2.7%) | 0.02 |
| Norovirus GI/GII | 50/485 (10%) | 27/268 (10%) | 0.01 | 23/234 (9.8%) | 15/158 (9.5%) | 0.01 | 27/251 (11%) | 12/110 (11%) | 0.00 |
| Rotavirus A | 5/485 (1%) | 5/268 (1.9%) | 0.07 | 1/234 (0.43%) | 2/158 (1.3%) | 0.09 | 4/251 (1.6%) | 3/110 (2.7%) | 0.08 |
| Coinfection, ≥2 GPP infections | 251/485 (52%) | 140/268 (52%) | 0.01 | 126/234 (54%) | 80/158 (51%) | 0.06 | 125/251 (50%) | 60/110 (55%) | 0.10 |
| Any STH infection | 202/447 (45%) | 106/242 (44%) | 0.03 | 106/218 (49%) | 64/142 (45%) | 0.07 | 96/229 (42%) | 42/100 (42%) | 0.00 |
| *Ascaris* | 109/447 (24%) | 54/242 (22%) | 0.05 | 65/218 (30%) | 30/142 (21%) | 0.20 | 44/229 (19%) | 24/100 (24%) | 0.12 |
| *Trichuris* | 170/447 (38%) | 86/242 (36%) | 0.05 | 85/218 (39%) | 54/142 (38%) | 0.02 | 85/229 (37%) | 32/100 (32%) | 0.11 |
| Coinfection, ≥2 STH infections | 77/447 (17%) | 34/242 (14%) | 0.09 | 44/218 (20%) | 20/142 (14%) | 0.16 | 33/229 (14%) | 14/100 (14%) | 0.01 |
| Number of GPP infections | 1.6 (1.1) | 1.7 (1.1) | 0.07 | 1.6 (1.1) | 1.6 (1.1) | 0.02 | 1.6 (1.1) | 1.7 (1.2) | 0.14 |
| Number of STH infections | 0.64 (0.77) | 0.58 (0.73) | 0.08 | 0.7 (0.79) | 0.59 (0.73) | 0.14 | 0.59 (0.75) | 0.57 (0.73) | 0.03 |
| **Child-, household-, compound-level characteristics** | | | | | | | | | |
| Child sex, female | 319/614 (52%) | 174/350 (50%) | 0.04 | 169/312 (54%) | 97/208 (47%) | 0.15 | 150/302 (50%) | 77/142 (54%) | 0.09 |
| Child breastfed | 206/609 (34%) | 106/365 (29%) | 0.10 | 107/310 (35%) | 62/216 (29%) | 0.13 | 99/299 (33%) | 44/149 (30%) | 0.08 |
| Child exclusively breastfed | 51/609 (8.4%) | 35/365 (9.6%) | 0.04 | 27/310 (8.7%) | 22/216 (10%) | 0.05 | 24/299 (8%) | 13/149 (8.7%) | 0.03 |
| Child age at survey, days | 697 (409) | 697 (396) | 0.00 | 698 (409) | 703 (400) | 0.01 | 696 (409) | 689 (391) | 0.02 |
| Child age at sampling, days | 668 (399) | 656 (382) | 0.03 | 661 (397) | 655 (395) | 0.02 | 674 (402) | 657 (364) | 0.04 |
| Child wears diapers | 402/609 (66%) | 234/364 (64%) | 0.04 | 209/310 (67%) | 133/216 (62%) | 0.12 | 193/299 (65%) | 101/148 (68%) | 0.08 |
| Child feces disposed in latrine | 173/609 (28%) | 116/365 (32%) | 0.07 | 79/310 (25%) | 69/216 (32%) | 0.14 | 94/299 (31%) | 47/149 (32%) | 0.00 |
| Caregiver completed primary school | 333/614 (54%) | 193/365 (53%) | 0.03 | 163/312 (52%) | 124/216 (57%) | 0.10 | 170/302 (56%) | 69/149 (46%) | 0.20 |
| Mother alive | 576/590 (98%) | 353/358 (99%) | 0.07 | 295/301 (98%) | 208/212 (98%) | 0.01 | 281/289 (97%) | 145/146 (99%) | 0.16 |
| Respondent is child's mother | 414/605 (68%) | 238/357 (67%) | 0.04 | 222/307 (72%) | 146/212 (69%) | 0.08 | 192/298 (64%) | 92/145 (63%) | 0.02 |
| Household floors covered | 575/615 (94%) | 349/368 (95%) | 0.06 | 300/313 (96%) | 211/217 (97%) | 0.08 | 275/302 (91%) | 138/151 (91%) | 0.01 |
| Household walls made of sturdy material | 399/615 (65%) | 243/368 (66%) | 0.02 | 216/313 (69%) | 154/217 (71%) | 0.04 | 183/302 (61%) | 89/151 (59%) | 0.03 |
| Latrine has drop-hole | 359/604 (59%) | 193/364 (53%) | 0.13 | 169/307 (55%) | 109/214 (51%) | 0.08 | 190/297 (64%) | 84/150 (56%) | 0.16 |
| Latrine has vent-pipe | 93/605 (15%) | 44/364 (12%) | 0.10 | 21/308 (6.8%) | 12/214 (5.6%) | 0.05 | 72/297 (24%) | 32/150 (21%) | 0.07 |
| Latrine has ceramic or concrete slab or pedestal | 224/602 (37%) | 133/363 (37%) | 0.01 | 101/305 (33%) | 80/213 (38%) | 0.09 | 123/297 (41%) | 53/150 (35%) | 0.13 |
| Latrine has sturdy walls | 193/605 (32%) | 110/363 (30%) | 0.03 | 84/306 (27%) | 58/215 (27%) | 0.01 | 109/299 (36%) | 52/148 (35%) | 0.03 |
| Water tap on compound grounds | 468/606 (77%) | 285/364 (78%) | 0.03 | 224/308 (73%) | 162/214 (76%) | 0.07 | 244/298 (82%) | 123/150 (82%) | 0.00 |
| Household crowding, ≥3 persons/room | 122/615 (20%) | 45/368 (12%) | 0.21 | 55/313 (18%) | 22/217 (10%) | 0.22 | 67/302 (22%) | 23/151 (15%) | 0.18 |
| Compound electricity normally functions | 556/615 (90%) | 331/372 (89%) | 0.05 | 272/313 (87%) | 195/220 (89%) | 0.05 | 284/302 (94%) | 136/152 (89%) | 0.17 |
| Standing water observed in compound | 44/605 (7.3%) | 26/363 (7.2%) | 0.00 | 7/306 (2.3%) | 7/215 (3.3%) | 0.06 | 37/299 (12%) | 19/148 (13%) | 0.01 |
| Leaking or standing wastewater observed in compound | 371/605 (61%) | 233/363 (64%) | 0.06 | 214/306 (70%) | 149/215 (69%) | 0.01 | 157/299 (53%) | 84/148 (57%) | 0.09 |
| Any animal observed | 395/615 (64%) | 226/372 (61%) | 0.07 | 189/313 (60%) | 129/220 (59%) | 0.04 | 206/302 (68%) | 97/152 (64%) | 0.09 |
| Dog observed | 51/615 (8.3%) | 23/372 (6.2%) | 0.08 | 18/313 (5.8%) | 10/220 (4.5%) | 0.05 | 33/302 (11%) | 13/152 (8.6%) | 0.08 |
| Chicken or duck observed | 94/615 (15%) | 36/372 (9.7%) | 0.17 | 43/313 (14%) | 27/220 (12%) | 0.04 | 51/302 (17%) | 9/152 (5.9%) | 0.35 |
| Cat observed | 341/615 (55%) | 205/372 (55%) | 0.01 | 167/313 (53%) | 120/220 (55%) | 0.02 | 174/302 (58%) | 85/152 (56%) | 0.03 |
| Faeces or used diapers observed around compound | 276/605 (46%) | 177/363 (49%) | 0.06 | 166/306 (54%) | 116/215 (54%) | 0.01 | 110/299 (37%) | 61/148 (41%) | 0.09 |
| Compound floods during rain | 377/615 (61%) | 226/372 (61%) | 0.01 | 211/313 (67%) | 137/220 (62%) | 0.11 | 166/302 (55%) | 89/152 (59%) | 0.07 |
| Number of household members | 6.4 (3.3) | 5.6 (2.6) | 0.27 | 6 (3) | 5.2 (2.1) | 0.33 | 6.8 (3.5) | 6.3 (3.1) | 0.18 |
| Household wealth score, 0-1 | 0.43 (0.1) | 0.44 (0.099) | 0.10 | 0.44 (0.1) | 0.45 (0.097) | 0.15 | 0.43 (0.1) | 0.43 (0.1) | 0.01 |
| Number of households in compound | 5.2 (4.6) | 4.7 (4.4) | 0.11 | 4.4 (2.9) | 3.8 (1.7) | 0.21 | 6.1 (5.6) | 6 (6.4) | 0.02 |
| Compound population | 21 (15) | 19 (14) | 0.18 | 17 (8.1) | 15 (6.1) | 0.22 | 26 (18) | 24 (20) | 0.11 |
| Number of water taps in compound | 1.5 (2.2) | 1.2 (1) | 0.22 | 1 (1.1) | 0.97 (0.83) | 0.04 | 2.1 (2.8) | 1.4 (1.2) | 0.30 |
| Number of latrines/drop-holes in compound | 1.1 (0.63) | 1.1 (0.65) | 0.00 | 1 (0.24) | 1 (0.2) | 0.04 | 1.2 (0.86) | 1.3 (0.97) | 0.03 |
| Compound population density | 0.084 (0.046) | 0.078 (0.045) | 0.13 | 0.076 (0.04) | 0.07 (0.039) | 0.14 | 0.092 (0.051) | 0.089 (0.05) | 0.06 |

Results are presented as prevalence (n/N (%)) or mean (standard deviation) at baseline. * Prevalence (or mean (SD)) for children with repeated observations at baseline and 12-month visits. † Prevalence (or mean (SD)) for children with observations at baseline visit and not the 12-month visit. ‡ Standardized mean difference between observations of children with and without repeated measures at baseline and 12-month visits. ⁑ Could not be calculated. Source files available in Supplemental Table 3 - source data 1 and Supplemental Table 3 - source code 1.

Supplemental Table 4: Baseline enrollment characteristics of children with and without repeated measures at the 24-month phase. Results are presented for all children combined and stratified by study arm.

|  | **All children** | | | **Control** | | | **Intervention** | | |
| --- | --- | --- | --- | --- | --- | --- | --- | --- | --- |
|  | **BL & 24M*** | **BL only†** | **Std. Diff.‡** | **BL & 24M** | **BL only** | **Std. Diff.** | **BL & 24M** | **BL only** | **Std. Diff.** |
| **Outcomes** | | | | | | | | | |
| Diarrhea | 75/504 (15%) | 51/470 (11%) | 0.12 | 35/244 (14%) | 32/282 (11%) | 0.09 | 40/260 (15%) | 19/188 (10%) | 0.16 |
| Any bacterial or protozoan infection | 310/394 (79%) | 281/359 (78%) | 0.01 | 144/183 (79%) | 169/209 (81%) | 0.05 | 166/211 (79%) | 112/150 (75%) | 0.09 |
| Any GPP infection | 322/394 (82%) | 293/359 (82%) | 0.00 | 148/183 (81%) | 175/209 (84%) | 0.07 | 174/211 (82%) | 118/150 (79%) | 0.10 |
| Any bacterial infection | 251/394 (64%) | 247/359 (69%) | 0.11 | 120/183 (66%) | 151/209 (72%) | 0.14 | 131/211 (62%) | 96/150 (64%) | 0.04 |
| *Shigella* | 158/394 (40%) | 173/359 (48%) | 0.16 | 74/183 (40%) | 105/209 (50%) | 0.20 | 84/211 (40%) | 68/150 (45%) | 0.11 |
| ETEC | 115/394 (29%) | 111/359 (31%) | 0.04 | 53/183 (29%) | 63/209 (30%) | 0.03 | 62/211 (29%) | 48/150 (32%) | 0.06 |
| *Campylobacter* | 31/394 (7.9%) | 29/359 (8.1%) | 0.01 | 18/183 (9.8%) | 21/209 (10%) | 0.01 | 13/211 (6.2%) | 8/150 (5.3%) | 0.04 |
| *C. difficile* | 18/394 (4.6%) | 17/359 (4.7%) | 0.01 | 10/183 (5.5%) | 12/209 (5.7%) | 0.01 | 8/211 (3.8%) | 5/150 (3.3%) | 0.02 |
| *E. coli* O157 | 17/394 (4.3%) | 14/359 (3.9%) | 0.02 | 7/183 (3.8%) | 6/209 (2.9%) | 0.05 | 10/211 (4.7%) | 8/150 (5.3%) | 0.03 |
| STEC | 6/394 (1.5%) | 7/359 (1.9%) | 0.03 | 2/183 (1.1%) | 1/209 (0.48%) | 0.07 | 4/211 (1.9%) | 6/150 (4%) | 0.12 |
| Any protozoan infection | 214/394 (54%) | 186/359 (52%) | 0.05 | 96/183 (52%) | 109/209 (52%) | 0.01 | 118/211 (56%) | 77/150 (51%) | 0.09 |
| *Giardia* | 204/394 (52%) | 183/359 (51%) | 0.02 | 92/183 (50%) | 109/209 (52%) | 0.04 | 112/211 (53%) | 74/150 (49%) | 0.08 |
| *Cryptosporidium* | 20/394 (5.1%) | 4/359 (1.1%) | 0.23 | 7/183 (3.8%) | 1/209 (0.48%) | 0.23 | 13/211 (6.2%) | 3/150 (2%) | 0.21 |
| *E. histolytica* | 2/394 (0.51%) | 2/359 (0.56%) | 0.01 | 0/183 (0.0%) | 0/209 (0.0%) | ..⁑ | 2/211 (0.95%) | 2/150 (1.3%) | 0.04 |
| Any viral infection | 55/394 (14%) | 50/359 (14%) | 0.00 | 22/183 (12%) | 31/209 (15%) | 0.08 | 33/211 (16%) | 19/150 (13%) | 0.09 |
| Adenovirus 40/41 | 14/394 (3.5%) | 8/359 (2.2%) | 0.08 | 7/183 (3.8%) | 6/209 (2.9%) | 0.05 | 7/211 (3.3%) | 2/150 (1.3%) | 0.13 |
| Norovirus GI/GII | 42/394 (11%) | 35/359 (9.8%) | 0.03 | 15/183 (8.2%) | 23/209 (11%) | 0.10 | 27/211 (13%) | 12/150 (8%) | 0.16 |
| Rotavirus A | 3/394 (0.76%) | 7/359 (1.9%) | 0.10 | 1/183 (0.55%) | 2/209 (0.96%) | 0.05 | 2/211 (0.95%) | 5/150 (3.3%) | 0.17 |
| Coinfection, ≥2 GPP infections | 206/394 (52%) | 185/359 (52%) | 0.02 | 97/183 (53%) | 109/209 (52%) | 0.02 | 109/211 (52%) | 76/150 (51%) | 0.02 |
| Any STH infection | 156/362 (43%) | 152/327 (46%) | 0.07 | 80/171 (47%) | 90/189 (48%) | 0.02 | 76/191 (40%) | 62/138 (45%) | 0.10 |
| *Ascaris* | 85/362 (23%) | 78/327 (24%) | 0.01 | 50/171 (29%) | 45/189 (24%) | 0.12 | 35/191 (18%) | 33/138 (24%) | 0.14 |
| *Trichuris* | 128/362 (35%) | 128/327 (39%) | 0.08 | 63/171 (37%) | 76/189 (40%) | 0.07 | 65/191 (34%) | 52/138 (38%) | 0.08 |
| Coinfection, ≥2 STH infections | 57/362 (16%) | 54/327 (17%) | 0.02 | 33/171 (19%) | 31/189 (16%) | 0.08 | 24/191 (13%) | 23/138 (17%) | 0.12 |
| Number of GPP infections | 1.6 (1.1) | 1.6 (1.2) | 0.04 | 1.6 (1.1) | 1.7 (1.1) | 0.10 | 1.6 (1.1) | 1.6 (1.2) | 0.01 |
| Number of STH infections | 0.61 (0.75) | 0.64 (0.76) | 0.04 | 0.67 (0.78) | 0.65 (0.75) | 0.03 | 0.55 (0.72) | 0.63 (0.77) | 0.10 |
| **Child-, household-, compound-level characteristics** | | | | | | | | | |
| Child sex, female | 260/503 (52%) | 233/461 (51%) | 0.02 | 124/241 (51%) | 142/279 (51%) | 0.01 | 136/262 (52%) | 91/182 (50%) | 0.04 |
| Child breastfed | 172/504 (34%) | 140/470 (30%) | 0.09 | 87/244 (36%) | 82/282 (29%) | 0.14 | 85/260 (33%) | 58/188 (31%) | 0.04 |
| Child exclusively breastfed | 35/504 (6.9%) | 51/470 (11%) | 0.14 | 19/244 (7.8%) | 30/282 (11%) | 0.10 | 16/260 (6.2%) | 21/188 (11%) | 0.18 |
| Child age at survey, days | 698 (403) | 696 (405) | 0.01 | 689 (400) | 709 (410) | 0.05 | 707 (406) | 675 (398) | 0.08 |
| Child age at sampling, days | 675 (406) | 651 (379) | 0.06 | 666 (403) | 652 (390) | 0.04 | 682 (409) | 650 (364) | 0.08 |
| Child wears diapers | 343/504 (68%) | 293/469 (62%) | 0.12 | 171/244 (70%) | 171/282 (61%) | 0.20 | 172/260 (66%) | 122/187 (65%) | 0.02 |
| Child feces disposed in latrine | 138/504 (27%) | 151/470 (32%) | 0.10 | 57/244 (23%) | 91/282 (32%) | 0.20 | 81/260 (31%) | 60/188 (32%) | 0.02 |
| Caregiver completed primary school | 274/507 (54%) | 252/472 (53%) | 0.01 | 131/245 (53%) | 156/283 (55%) | 0.03 | 143/262 (55%) | 96/189 (51%) | 0.08 |
| Mother alive | 474/486 (98%) | 455/462 (98%) | 0.07 | 232/236 (98%) | 271/277 (98%) | 0.03 | 242/250 (97%) | 184/185 (99%) | 0.20 |
| Respondent is child's mother | 337/500 (67%) | 315/462 (68%) | 0.02 | 173/241 (72%) | 195/278 (70%) | 0.04 | 164/259 (63%) | 120/184 (65%) | 0.04 |
| Household floors covered | 469/507 (93%) | 455/476 (96%) | 0.13 | 233/245 (95%) | 278/285 (98%) | 0.13 | 236/262 (90%) | 177/191 (93%) | 0.09 |
| Household walls made of sturdy material | 337/507 (66%) | 305/476 (64%) | 0.05 | 184/245 (75%) | 186/285 (65%) | 0.22 | 153/262 (58%) | 119/191 (62%) | 0.08 |
| Latrine has drop-hole | 294/497 (59%) | 258/471 (55%) | 0.09 | 133/239 (56%) | 145/282 (51%) | 0.08 | 161/258 (62%) | 113/189 (60%) | 0.05 |
| Latrine has vent-pipe | 80/497 (16%) | 57/472 (12%) | 0.12 | 18/239 (7.5%) | 15/283 (5.3%) | 0.09 | 62/258 (24%) | 42/189 (22%) | 0.04 |
| Latrine has ceramic or concrete slab or pedestal | 184/494 (37%) | 173/471 (37%) | 0.01 | 77/236 (33%) | 104/282 (37%) | 0.09 | 107/258 (41%) | 69/189 (37%) | 0.10 |
| Latrine has sturdy walls | 165/501 (33%) | 138/467 (30%) | 0.07 | 67/240 (28%) | 75/281 (27%) | 0.03 | 98/261 (38%) | 63/186 (34%) | 0.08 |
| Water tap on compound grounds | 389/498 (78%) | 364/472 (77%) | 0.02 | 171/239 (72%) | 215/283 (76%) | 0.10 | 218/259 (84%) | 149/189 (79%) | 0.14 |
| Household crowding, ≥3 persons/room | 114/507 (22%) | 53/476 (11%) | 0.31 | 45/245 (18%) | 32/285 (11%) | 0.20 | 69/262 (26%) | 21/191 (11%) | 0.40 |
| Compound electricity normally functions | 454/507 (90%) | 433/480 (90%) | 0.02 | 214/245 (87%) | 253/288 (88%) | 0.02 | 240/262 (92%) | 180/192 (94%) | 0.08 |
| Standing water observed in compound | 39/501 (7.8%) | 31/467 (6.6%) | 0.04 | 7/240 (2.9%) | 7/281 (2.5%) | 0.03 | 32/261 (12%) | 24/186 (13%) | 0.02 |
| Leaking or standing wastewater observed in compound | 308/501 (61%) | 296/467 (63%) | 0.04 | 164/240 (68%) | 199/281 (71%) | 0.05 | 144/261 (55%) | 97/186 (52%) | 0.06 |
| Any animal observed | 337/507 (66%) | 284/480 (59%) | 0.15 | 156/245 (64%) | 162/288 (56%) | 0.15 | 181/262 (69%) | 122/192 (64%) | 0.12 |
| Dog observed | 49/507 (9.7%) | 25/480 (5.2%) | 0.17 | 17/245 (6.9%) | 11/288 (3.8%) | 0.14 | 32/262 (12%) | 14/192 (7.3%) | 0.17 |
| Chicken or duck observed | 71/507 (14%) | 59/480 (12%) | 0.05 | 32/245 (13%) | 38/288 (13%) | 0.00 | 39/262 (15%) | 21/192 (11%) | 0.12 |
| Cat observed | 294/507 (58%) | 252/480 (53%) | 0.11 | 143/245 (58%) | 144/288 (50%) | 0.17 | 151/262 (58%) | 108/192 (56%) | 0.03 |
| Feces or used diapers observed around compound | 218/501 (44%) | 235/467 (50%) | 0.14 | 120/240 (50%) | 162/281 (58%) | 0.15 | 98/261 (38%) | 73/186 (39%) | 0.03 |
| Compound floods during rain | 310/507 (61%) | 293/480 (61%) | 0.00 | 166/245 (68%) | 182/288 (63%) | 0.10 | 144/262 (55%) | 111/192 (58%) | 0.06 |
| Number of household members | 6.7 (3.4) | 5.5 (2.6) | 0.39 | 6.3 (3) | 5.2 (2.2) | 0.42 | 7.1 (3.6) | 6.1 (3) | 0.31 |
| Household wealth score, 0-1 | 0.43 (0.11) | 0.44 (0.097) | 0.12 | 0.44 (0.1) | 0.45 (0.095) | 0.10 | 0.42 (0.11) | 0.43 (0.1) | 0.11 |
| Number of households in compound | 5.3 (4.7) | 4.7 (4.3) | 0.13 | 4.4 (3.1) | 3.9 (1.8) | 0.21 | 6.1 (5.7) | 5.9 (6.2) | 0.03 |
| Compound population | 22 (15) | 18 (14) | 0.26 | 17 (8.1) | 15 (6.5) | 0.27 | 27 (18) | 23 (19) | 0.18 |
| Number of water taps in compound | 1.6 (2.2) | 1.2 (1.3) | 0.24 | 1 (1) | 0.99 (0.92) | 0.02 | 2.2 (2.8) | 1.4 (1.8) | 0.31 |
| Number of latrines in compound | 1.1 (0.62) | 1.1 (0.65) | 0.01 | 1 (0.25) | 1 (0.19) | 0.04 | 1.2 (0.82) | 1.3 (0.99) | 0.08 |
| Compound population density | 0.084 (0.049) | 0.079 (0.042) | 0.13 | 0.072 (0.038) | 0.075 (0.04) | 0.05 | 0.096 (0.055) | 0.084 (0.044) | 0.23 |

Results are presented as prevalence (n/N (%)) or mean (standard deviation) at baseline. * Prevalence (or mean (SD)) for children with repeated observations at baseline and 24-month visits. † Prevalence (or mean (SD)) for children with observations at the baseline visit and not the 24-month visit. ‡ Standardized mean difference between observations of children with and without repeated measures at baseline and 24-month visits. ⁑ Could not be calculated. Source files available in Supplemental Table 4 - source data 1 and Supplemental Table 4 - source code 1.

### Supplemental Table 5: Balance of characteristics measured at 12-month visits between children with repeat observations at baseline and 12-month and children with observations at the 12-month phase only.

|  | **All Children** | | | **Control** | | | **Intervention** | | |  |
| --- | --- | --- | --- | --- | --- | --- | --- | --- | --- | --- |
|  | **BL & 12M*** | **12M only†** | **Std. Diff.‡** | **BL & 12M** | **12M only** | **Std. Diff.** | **BL & 12M** | **12M only** | **Std. Diff.** | **Std. Diff. Control v. Interv.⁑** |
| Child sex, female | 319/614 (52%) | 156/313 (50%) | 0.04 | 169/312 (54%) | 73/155 (47%) | 0.14 | 150/302 (50%) | 83/158 (53%) | 0.06 | 0.11 |
| Child breastfed | 27/562 (4.8%) | 161/305 (53%) | 1.25 | 13/280 (4.6%) | 76/151 (50%) | 1.19 | 14/282 (5%) | 85/154 (55%) | 1.31 | 0.10 |
| Child exclusively breastfed | 3/562 (0.53%) | 38/305 (12%) | 0.50 | 2/280 (0.71%) | 16/151 (11%) | 0.44 | 1/282 (0.35%) | 22/154 (14%) | 0.56 | 0.11 |
| Caregiver completed primary school | 305/614 (50%) | 144/309 (47%) | 0.06 | 156/312 (50%) | 62/153 (41%) | 0.19 | 149/302 (49%) | 82/156 (53%) | 0.06 | 0.24 |
| Child wears diapers | 83/563 (15%) | 194/305 (64%) | 1.16 | 40/281 (14%) | 92/151 (61%) | 1.10 | 43/282 (15%) | 102/154 (66%) | 1.21 | 0.11 |
| Respondent is child's mother | 365/563 (65%) | 236/305 (77%) | 0.28 | 188/281 (67%) | 121/151 (80%) | 0.30 | 177/282 (63%) | 115/154 (75%) | 0.26 | 0.13 |
| Household floors covered | 584/615 (95%) | 305/321 (95%) | 0.00 | 299/313 (96%) | 155/163 (95%) | 0.02 | 285/302 (94%) | 150/158 (95%) | 0.03 | 0.01 |
| Household walls made of sturdy material | 398/615 (65%) | 189/321 (59%) | 0.12 | 212/313 (68%) | 101/163 (62%) | 0.12 | 186/302 (62%) | 88/158 (56%) | 0.12 | 0.13 |
| Household crowding, ≥3 persons/room | 210/615 (34%) | 106/321 (33%) | 0.02 | 111/313 (35%) | 54/163 (33%) | 0.05 | 99/302 (33%) | 52/158 (33%) | 0.00 | 0.00 |
| Compound electricity normally functions | 575/615 (94%) | 304/324 (94%) | 0.01 | 286/313 (91%) | 152/164 (93%) | 0.05 | 289/302 (96%) | 152/160 (95%) | 0.03 | 0.10 |
| Any animal observed | 505/611 (83%) | 275/324 (85%) | 0.06 | 235/309 (76%) | 131/164 (80%) | 0.09 | 270/302 (89%) | 144/160 (90%) | 0.02 | 0.29 |
| Dog observed | 134/611 (22%) | 81/324 (25%) | 0.07 | 57/309 (18%) | 37/164 (23%) | 0.10 | 77/302 (26%) | 44/160 (28%) | 0.05 | 0.11 |
| Chicken or duck observed | 77/611 (13%) | 42/324 (13%) | 0.01 | 34/309 (11%) | 18/164 (11%) | 0.00 | 43/302 (14%) | 24/160 (15%) | 0.02 | 0.12 |
| Cat observed | 469/611 (77%) | 249/324 (77%) | 0.00 | 218/309 (71%) | 118/164 (72%) | 0.03 | 251/302 (83%) | 131/160 (82%) | 0.03 | 0.24 |
| Compound floods during rain | 220/615 (36%) | 119/324 (37%) | 0.02 | 132/313 (42%) | 64/164 (39%) | 0.06 | 88/302 (29%) | 55/160 (34%) | 0.11 | 0.10 |
| Child age at survey, days | 1114 (415) | 622 (502) | 1.07 | 1105 (413) | 684 (535) | 0.88 | 1122 (417) | 560 (461) | 1.28 | 0.25 |
| Child age at sampling, days | 1102 (417) | 605 (484) | 1.10 | 1080 (414) | 649 (516) | 0.92 | 1122 (420) | 563 (450) | 1.29 | 0.18 |
| Number of household members | 6.5 (3.2) | 6.3 (3.3) | 0.06 | 6.2 (3) | 6.4 (3.5) | 0.05 | 6.8 (3.3) | 6.2 (3.2) | 0.17 | 0.05 |
| Household wealth score, 0-1 | 0.4 (0.11) | 0.39 (0.11) | 0.02 | 0.4 (0.11) | 0.39 (0.11) | 0.12 | 0.39 (0.1) | 0.4 (0.1) | 0.10 | 0.11 |
| Number of households in compound | 5.2 (4.7) | 5.4 (5.5) | 0.04 | 4.2 (2.9) | 4 (2.3) | 0.09 | 6.3 (5.9) | 6.9 (7.3) | 0.09 | 0.53 |
| Compound population | 23 (22) | 24 (26) | 0.04 | 18 (9.7) | 18 (8.7) | 0.05 | 28 (29) | 30 (35) | 0.07 | 0.50 |
| Compound population density | 0.086 (0.049) | 0.084 (0.051) | 0.04 | 0.08 (0.043) | 0.078 (0.044) | 0.05 | 0.091 (0.054) | 0.089 (0.058) | 0.03 | 0.22 |

Results are presented as prevalence (n/N (%)) or mean (standard deviation) at 12-month visit. * Prevalence (or mean (SD)) for children with repeated observations at baseline and 12-month visits. † Prevalence (or mean (SD)) for children with observations at the 12-month visit only. ‡ Standardized mean difference between observations of children with and without repeated measures at baseline and 12-month visits. ⁑ Standardized mean difference between observations from control and intervention children measured at 12-month visit only. Source files available in Supplemental Table 5 - source data 1 and Supplemental Table 5 - source code 1.

### Supplemental Table 6: Balance of characteristics measured at 24-month visits between children with repeat observations at baseline and 24-month and children with observations at the 24-month phase only.

|  | **All Children** | | | **Control** | | | **Intervention** | | |  |
| --- | --- | --- | --- | --- | --- | --- | --- | --- | --- | --- |
|  | **BL & 24M*** | **24M only†** | **Std. Diff.†** | **BL & 24M** | **24M only** | **Std. Diff.** | **BL & 24M** | **24M only** | **Std. Diff.** | **Std. Diff. Control v. Interv.⁑** |
| Child sex, female | 260/503 (52%) | 190/428 (44%) | 0.15 | 124/241 (51%) | 96/222 (43%) | 0.16 | 136/262 (52%) | 94/206 (46%) | 0.13 | 0.05 |
| Child breastfed | 0/418 (0.0% | 129/381 (34%) | 1.01 | 0/195 (0.0%) | 68/194 (35%) | 1.04 | 0/223 (0.0%) | 61/187 (33%) | 0.98 | 0.05 |
| Child exclusively breastfed | 0/418 (0.0%) | 36/381 (9.4%) | 0.46 | 0/195 (0.0%) | 16/194 (8.3%) | 0.42 | 0/223 (0.0%) | 20/187 (11%) | 0.49 | 0.08 |
| Caregiver completed primary school | 199/507 (39%) | 164/427 (38%) | 0.02 | 88/245 (36%) | 82/221 (37%) | 0.02 | 111/262 (42%) | 82/206 (40%) | 0.05 | 0.06 |
| Child wears diapers | 3/419 (0.72%) | 196/381 (51%) | 1.42 | 1/196 (0.51%) | 101/194 (52%) | 1.44 | 2/223 (0.9%) | 95/187 (51%) | 1.39 | 0.03 |
| Respondent is child's mother | 259/419 (62%) | 298/381 (78%) | 0.36 | 129/196 (66%) | 161/194 (83%) | 0.40 | 130/223 (58%) | 137/187 (73%) | 0.32 | 0.24 |
| Household floors covered | 484/507 (95%) | 459/467 (98%) | 0.16 | 237/245 (97%) | 234/239 (98%) | 0.07 | 247/262 (94%) | 225/228 (99%) | 0.24 | 0.06 |
| Household walls made of sturdy material | 352/507 (69%) | 296/467 (63%) | 0.13 | 180/245 (73%) | 157/239 (66%) | 0.17 | 172/262 (66%) | 139/228 (61%) | 0.10 | 0.10 |
| Household crowding, ≥3 persons/room | 137/507 (27%) | 108/467 (23%) | 0.09 | 74/245 (30%) | 66/239 (28%) | 0.06 | 63/262 (24%) | 42/228 (18%) | 0.14 | 0.22 |
| Compound electricity normally functions | 485/507 (96%) | 472/494 (96%) | 0.01 | 230/245 (94%) | 237/254 (93%) | 0.02 | 255/262 (97%) | 235/240 (98%) | 0.04 | 0.23 |
| Any animal observed | 384/507 (76%) | 359/494 (73%) | 0.07 | 162/245 (66%) | 182/254 (72%) | 0.12 | 222/262 (85%) | 177/240 (74%) | 0.27 | 0.05 |
| Dog observed | 70/507 (14%) | 78/494 (16%) | 0.06 | 30/245 (12%) | 40/254 (16%) | 0.10 | 40/262 (15%) | 38/240 (16%) | 0.02 | 0.00 |
| Chicken or duck observed | 63/507 (12%) | 52/494 (11%) | 0.06 | 22/245 (9%) | 32/254 (13%) | 0.12 | 41/262 (16%) | 20/240 (8.3%) | 0.23 | 0.14 |
| Cat observed | 360/507 (71%) | 340/494 (69%) | 0.05 | 154/245 (63%) | 174/254 (69%) | 0.12 | 206/262 (79%) | 166/240 (69%) | 0.22 | 0.01 |
| Compound floods during rain | 182/507 (36%) | 184/494 (37%) | 0.03 | 89/245 (36%) | 107/254 (42%) | 0.12 | 93/262 (36%) | 77/240 (32%) | 0.07 | 0.21 |
| Child age at survey, days | 1518 (407) | 740 (518) | 1.67 | 1520 (406) | 749 (541) | 1.61 | 1516 (408) | 731 (494) | 1.73 | 0.04 |
| Child age at sampling, days | 1510 (415) | 694 (478) | 1.82 | 1505 (408) | 716 (512) | 1.70 | 1516 (422) | 672 (439) | 1.96 | 0.09 |
| Number of household members | 6.6 (3.1) | 6.3 (3.4) | 0.10 | 6.5 (3) | 6.6 (3.8) | 0.04 | 6.7 (3.1) | 6 (2.8) | 0.26 | 0.20 |
| Household wealth score, 0-1 | 0.41 (0.11) | 0.41 (0.11) | 0.01 | 0.41 (0.12) | 0.4 (0.11) | 0.11 | 0.41 (0.1) | 0.42 (0.097) | 0.15 | 0.19 |
| Number of households in compound | 5.3 (4.9) | 5.5 (5.5) | 0.04 | 4.3 (2.8) | 4.4 (3.2) | 0.03 | 6.2 (6.1) | 6.6 (6.9) | 0.06 | 0.41 |
| Compound population | 21 (15) | 21 (16) | 0.04 | 18 (9.5) | 17 (8.9) | 0.07 | 25 (19) | 25 (21) | 0.00 | 0.47 |
| Compound population density | 0.08 (0.047) | 0.08 (0.047) | 0.01 | 0.074 (0.037) | 0.075 (0.042) | 0.03 | 0.087 (0.053) | 0.085 (0.052) | 0.03 | 0.22 |

Results are presented as prevalence (n/N (%)) or mean (standard deviation) at 24-month visit. * Prevalence (or mean (SD)) for children with repeated observations at baseline and 24-month visits. † Prevalence (or mean (SD)) for children with observations at the 24-month visit only. ‡ Standardized mean difference between observations of children with and without repeated measures at baseline and 24-month visits. ⁑ Standardized mean difference between observations from control and intervention children measured at 24-month visit only. Source files available in Supplemental Table 6 - source data 1 and Supplemental Table 6 - source code 1.

### Supplemental Table 7: Sensitivity analysis assessing the impact of reported deworming on STH effect estimates 12 and 24 months after the intervention.

|  | **12-month Prevalence ratio** | | | **24-month Prevalence ratio** | | |
| --- | --- | --- | --- | --- | --- | --- |
|  | **Main analysis, all children*** | **Adjusted for reported deworming †** | **Restricted to children dewormed at baseline ‡** | **Main analysis, all children*** | **Adjusted for reported deworming †** | **Adjusted for time since deworming⁑** |
|  | n=1239 | n=1239 | n=1031 | n=1161 | n=1161 | N=1159 |
| Any STH infection | 1.11 (0.89 - 1.38) | 1.09 (0.87 - 1.35) | 1.06 (0.84 - 1.33) | 0.95 (0.77 - 1.17) | 0.93 (0.77 - 1.16) | 0.93 (0.75 – 1.14) |
| Trichuris | 1.01 (0.79 - 1.28) | 0.98 (0.77 - 1.24) | 0.96 (0.74 - 1.23) | 0.86 (0.67 - 1.10) | 0.85 (0.66 - 1.08) | 0.86 (0.67 – 1.09) |
| Ascaris | 1.33 (0.92 - 1.93) | 1.30 (0.90 - 1.88) | 1.30 (0.87 - 1.94) | 0.83 (0.54 - 1.27) | 0.84 (0.55 - 1.29) | 0.78 (0.51 – 1.18) |
| Coinfection, ≥2 STH | 1.17 (0.76 - 1.79) | 1.12 (0.73 - 1.71) | 1.16 (0.73 - 1.85) | 0.63 (0.37 - 1.07) | 0.63 (0.37 - 1.08) | 0.60 (0.35 – 1.03) |

All effect estimates are presented as prevalence ratios (ratio of ratios) with 95% confidence intervals and estimated using generalized estimating equations to fit Poisson regression models with robust standard errors. All models adjusted for child age, sex, caregiver education level, and household wealth. *Analysis includes all children regardless of caregiver-reported deworming status.  †Analysis is adjusted for reported deworming status. Effect estimates at 12-month are adjusted for baseline deworming confirmation, effect estimates at 24-month are adjusted for baseline and/or 12-month deworming confirmation. ‡Analysis is restricted to children whose caregivers confirmed baseline deworming. ⁑ Adjusted for time between 12-month deworming and 24-month sample collection, time broken into 3 intervals: 0-3 months, 4-6 months, and >6 months. The NDC performed 12-month deworming activities at the end of the 12-month phase instead of concurrent to 12-month sample collection resulting in some variation in the amount of time between 12-month deworming and 24-month sample collection among participants. All samples collected during 12-month phase were collected >6 months after deworming and no adjustment for time since deworming was made. Source files available in Supplemental Table 7 - source data 1 and Supplemental Table 7 - source code 1.

### Supplemental Table 8: Sensitivity analysis assessing impact of independent upgrading of control sanitation facilities on effect estimates.

|  | **12-month adjusted prevalence ratio** | | **24-month adjusted prevalence ratio** | |
| --- | --- | --- | --- | --- |
|  | **Main analysis, all children*** | **Excluding controls with upgraded sanitation†** | **Main analysis, all children*** | **Excluding controls with upgraded sanitation†** |
| Any bacterial or protozoan infection | 1.04 (0.94 – 1.15), n=1510 | 1.05 (0.95 – 1.16), n=1491 | 0.99 (0.91 – 1.09), n=1536 | 1.00 (0.91 – 1.10), n=1502 |
| Any STH infection | 1.11 (0.89 – 1.38), n=1239 | 1.11 (0.89 – 1.38), n=1225 | 0.95 (0.77 – 1.17), n=1161 | 0.94 (0.76 – 1.16), n=1148 |
| Diarrhea | 1.69 (0.89 – 3.21), n=1594 | 1.76 (0.91 – 3.39), n=1575 | 0.84 (0.47 – 1.51), n=1502 | 0.81 (0.45 – 1.48), n=1471 |

All effect estimates are presented as prevalence ratios (ratio of ratios) with 95% confidence intervals and estimated using generalized estimating equations to fit Poisson regression models with robust standard errors. All infection outcomes are adjusted for child age and sex, caregiver’s education, and household wealth index, and the diarrhea outcome is also adjusted for baseline presence of a drop-hole cover and reported use of a tap on compound grounds as primary drinking water source. * Results represent effect estimates for the main analyses which included control children irrespective of whether their latrines had been independently upgraded (results also presented in Table 2 in main text). **†** Results from sensitivity analyses which exclude control children living in compounds that independently upgraded their latrines to be similar to the intervention. Source files available in Supplemental Table 8 - source data 1 and Supplemental Table 8 - source code 1.

### Supplemental Table 9: Confounding assessment for primary outcome and both secondary outcomes (any STH, diarrhea) at 12-month.

|  | n/N (%) or mean (SD) at Baseline | | Std diff.^*^ | Primary outcome Unadjusted^†^ | Primary outcome Adjusted^‡^ | Any STH Unadjusted^†^ | Any STH Adjusted^‡^ | Diarrhea Unadjusted^†^ | Diarrhea Adjusted^‡^ |
| --- | --- | --- | --- | --- | --- | --- | --- | --- | --- |
| Variable | Control | Inter-vention. |  | Comparator PR: 1.04 (0.94 - 1.15) | Comparator aPR: 1.04 (0.94 - 1.15) | Comparator PR: 1.12 (0.89 - 1.40) | Comparator aPR: 1.11 (0.90 - 1.38) | Comparator PR: 1.41 (0.80 - 2.48) | Comparator aPR: 1.32 (0.75 - 2.33) |
| Female | 266/520 (51%) | 227/444 (51%) | 0.00 | 1.04 (0.94 - 1.15) | 1.04 (0.94 - 1.15) | 1.14 (0.91 - 1.42) | 1.11 (0.89 - 1.38) | 1.39 (0.79 - 2.46) | 1.32 (0.75 - 2.33) |
| Any breastfeeding | 169/526 (32%) | 143/448 (32%) | 0.00 | 1.05 (0.95 - 1.15) | 1.05 (0.95 - 1.15) | 1.11 (0.90 - 1.38) | 1.11 (0.90 - 1.38) | 1.39 (0.79 - 2.45) | 1.33 (0.75 - 2.35) |
| Caregiver completed primary school | 287/528 (54%) | 239/451 (53%) | 0.03 | 1.04 (0.94 - 1.15) | 1.04 (0.94 - 1.15) | 1.12 (0.90 - 1.41) | 1.11 (0.89 - 1.38) | 1.40 (0.80 - 2.48) | 1.32 (0.75 - 2.33) |
| Respondent is mother | 368/519 (71%) | 284/443 (64%) | 0.15 | 1.05 (0.95 - 1.16) | 1.04 (0.94 - 1.15) | 1.13 (0.90 - 1.42) | 1.11 (0.89 - 1.38) | 1.37 (0.78 - 2.42) | 1.29 (0.73 - 2.28) |
| Household floors covered | 511/530 (96%) | 413/453 (91%) | 0.22 | 1.04 (0.94 - 1.15) | 1.04 (0.94 - 1.15) | 1.12 (0.89 - 1.40) | 1.12 (0.90 - 1.39) | 1.39 (0.79 - 2.47) | 1.32 (0.74 - 2.34) |
| Household walls made of sturdy material | 370/530 (70%) | 272/453 (60%) | 0.21 | 1.04 (0.94 - 1.15) | 1.04 (0.94 - 1.15) | 1.12 (0.89 - 1.40) | 1.11 (0.89 - 1.38) | 1.41 (0.80 - 2.48) | 1.32 (0.75 - 2.33) |
| Drinking water source in compound | 386/522 (74%) | 367/448 (82%) | 0.19 | 1.03 (0.93 - 1.15) | 1.03 (0.93 - 1.14) | 1.08 (0.85 - 1.36) | 1.05 (0.83 - 1.33) | 1.65 (0.89 - 3.06) | 1.59 (0.85 - 2.95) |
| Faeces visible around compound grounds | 282/521 (54%) | 171/447 (38%) | 0.32 | 1.03 (0.93 - 1.13) | 1.03 (0.93 - 1.13) | 1.14 (0.91 - 1.43) | 1.12 (0.90 - 1.40) | 1.43 (0.81 - 2.54) | 1.35 (0.76 - 2.40) |
| Compound floods when it rains | 348/533 (65%) | 255/454 (56%) | 0.19 | 1.04 (0.94 - 1.15) | 1.04 (0.94 - 1.15) | 1.12 (0.89 - 1.40) | 1.11 (0.89 - 1.38) | 1.41 (0.80 - 2.49) | 1.32 (0.74 - 2.33) |
| Latrine drop-hole has cover | 278/521 (53%) | 274/447 (61%) | 0.16 | 1.04 (0.94 - 1.15) | 1.03 (0.93 - 1.15) | 1.11 (0.88 - 1.40) | 1.08 (0.85 - 1.36) | 1.74 (0.92 - 3.30) | 1.69 (0.89 - 3.20) |
| Latrine has ceramic/concrete slab or pedestal | 181/518 (35%) | 176/447 (39%) | 0.09 | 1.04 (0.94 - 1.15) | 1.04 (0.93 - 1.15) | 1.10 (0.87 - 1.39) | 1.07 (0.85 - 1.35) | 1.71 (0.90 - 3.24) | 1.65 (0.87 - 3.14) |
| Latrine walls made of sturdy material | 142/521 (27%) | 161/447 (36%) | 0.19 | 1.03 (0.93 - 1.14) | 1.03 (0.93 - 1.13) | 1.14 (0.91 - 1.43) | 1.12 (0.90 - 1.40) | 1.42 (0.80 - 2.51) | 1.33 (0.75 - 2.37) |
| Standing water observed around compound | 14/521 (2.7%) | 56/447 (13%) | 0.38 | 1.03 (0.93 - 1.14) | 1.03 (0.93 - 1.13) | 1.14 (0.91 - 1.42) | 1.12 (0.90 - 1.39) | 1.42 (0.80 - 2.51) | 1.34 (0.75 - 2.38) |
| Leaking or standing wastewater observed around grounds | 363/521 (70%) | 241/447 (54%) | 0.33 | 1.03 (0.93 - 1.14) | 1.03 (0.93 - 1.13) | 1.14 (0.91 - 1.43) | 1.12 (0.90 - 1.40) | 1.42 (0.80 - 2.51) | 1.34 (0.75 - 2.38) |
| Compound has electricity that normally functions | 467/533 (88%) | 420/454 (93%) | 0.16 | 1.04 (0.94 - 1.15) | 1.04 (0.94 - 1.15) | 1.11 (0.89 - 1.39) | 1.11 (0.89 - 1.38) | 1.41 (0.80 - 2.48) | 1.32 (0.75 - 2.34) |
| Any animal observed in compound | 318/533 (60%) | 303/454 (67%) | 0.15 | 1.04 (0.95 - 1.15) | 1.04 (0.95 - 1.15) | 1.13 (0.91 - 1.41) | 1.13 (0.91 - 1.40) | 1.39 (0.79 - 2.44) | 1.29 (0.73 - 2.28) |
| Dog observed | 28/533 (5.3%) | 46/454 (10%) | 0.18 | 1.05 (0.95 - 1.15) | 1.04 (0.95 - 1.15) | 1.13 (0.90 - 1.41) | 1.12 (0.90 - 1.39) | 1.38 (0.79 - 2.40) | 1.30 (0.75 - 2.27) |
| Chicken or duck observed | 70/533 (13%) | 60/454 (13%) | 0.00 | 1.05 (0.95 - 1.15) | 1.05 (0.95 - 1.16) | 1.12 (0.90 - 1.41) | 1.12 (0.90 - 1.40) | 1.37 (0.78 - 2.40) | 1.27 (0.72 - 2.23) |
| Cat observed | 287/533 (54%) | 259/454 (57%) | 0.06 | 1.05 (0.95 - 1.16) | 1.04 (0.95 - 1.15) | 1.14 (0.91 - 1.42) | 1.13 (0.91 - 1.41) | 1.39 (0.79 - 2.45) | 1.30 (0.74 - 2.29) |
| Compound density, terciles |  |  | 0.40 | 1.05 (0.95 – 1.16) | 1.05 (0.95 – 1.16) | 1.10 (0.88 - 1.38) | 1.10 (0.89 - 1.38) | 1.43 (0.81 - 2.50) | 1.32 (0.75 - 2.33) |
| 0 (least dense) | 199/519 (38%) | 120/447 (27%) | .. | .. | .. | .. | .. | .. | .. |
| 1 | 191/519 (37%) | 137/447 (31%) | .. | .. | .. | .. | .. | .. | .. |
| 2 (most dense) | 129/519 (25%) | 190/447 (43%) | .. | .. | .. | .. | .. | .. | .. |
| Child age at survey, days | 700 (405) | 694 (403) | 0.02 | .. | .. | .. | .. | 1.33 (0.76 - 2.34) | 1.32 (0.75 - 2.33) |
| Child age at sample, days | 659 (396) | 669 (391) | 0.03 | 1.04 (0.94 - 1.14) | 1.04 (0.94 - 1.15) | 1.09 (0.88 - 1.36) | 1.11 (0.89 - 1.38) | - | - |
| Cumulative monthly rainfall at survey, mm | 22 (23) | 23 (24) | 0.07 | .. | .. | .. | .. | 1.39 (0.79 - 2.44) | 1.30 (0.74 - 2.29) |
| Cumulative monthly rainfall at sample, mm | 25 (30) | 32 (38) | 0.19 | 1.04 (0.94 - 1.15) | 1.04 (0.95 - 1.15) | 1.13 (0.90 - 1.41) | 1.13 (0.91 - 1.40) | .. | .. |
| Survey collected during rainy season | 155/526 (29%) | 222/448 (50%) | 0.42 | .. | .. | .. | .. | 1.44 (0.81 – 2.54) | 1.34 (0.76 – 2.38) |
| Sample collected during rainy season | 136/409 (33%) | 183/370 (49%) | 0.33 | 1.05 (0.95 – 1.16) | 1.05 (0.95 – 1.16) | 1.12 (0.90 – 1.40) | 1.12 (0.90 – 1.39) | .. | .. |
| Wealth score | 0.44 (0.1) | 0.43 (0.1) | 0.16 | 1.04 (0.94 - 1.15) | 1.04 (0.94 - 1.15) | 1.12 (0.90 - 1.40) | 1.11 (0.89 - 1.38) | 1.39 (0.79 - 2.46) | 1.32 (0.75 - 2.33) |
| Number of household residents | 5.7 (2.7) | 6.6 (3.4) | 0.32 | 1.04 (0.94 - 1.15) | 1.04 (0.94 - 1.15) | 1.13 (0.90 - 1.41) | 1.12 (0.90 - 1.39) | 1.38 (0.78 - 2.44) | 1.31 (0.74 - 2.31) |
| Number of Compound residents | 16 (7.3) | 25 (19) | 0.64 | 1.04 (0.94 - 1.15) | 1.04 (0.94 - 1.15) | 1.10 (0.88 - 1.37) | 1.09 (0.88 - 1.35) | 1.39 (0.79 - 2.45) | 1.31 (0.74 - 2.32) |
| Number of households in compound | 4.1 (2.5) | 6.1 (5.9) | 0.42 | 1.04 (0.94 – 1.15) | 1.04 (0.94 – 1.15) | 1.11 (0.89 – 1.37) | 1.09 (0.88 – 1.36) | 1.40 (0.79 – 2.46) | 1.31 (0.74 – 2.32) |
| Number of compound latrines | 1.0 (0.22) | 1.2 (0.9) | 0.33 | 1.04 (0.94 - 1.15) | 1.04 (0.94 - 1.15) | 1.13 (0.91 - 1.40) | 1.12 (0.90 - 1.39) | 1.40 (0.79 - 2.47) | 1.33 (0.75 - 2.35) |
| Number of compound waterpoints | 0.99 (0.98) | 1.9 (2.4) | 0.47 | 1.03 (0.93 - 1.14) | 1.03 (0.93 - 1.14) | 1.13 (0.91 - 1.42) | 1.12 (0.90 - 1.39) | 1.45 (0.82 - 2.56) | 1.37 (0.77 - 2.43) |

*Standardized difference between arms in baseline covariates. † Compared with 12-month unadjusted prevalence ratio (12-month difference-in-difference estimator). ‡ Compared with 12-month prevalence ratio adjusted for *a priori* covariates child age, sex, caregiver education, and poverty (wealth score). Source files available in Supplemental Table 9 - source data 1 and Supplemental Table 9 - source code 1.

### Supplemental Table 10: Effect estimates (prevalence ratios) for main analyses and all sub-group analyses adjusted for *a priori* covariates and age-squared

|  | Main analysis, all children† | | Sub-group analysis, children born after intervention* | | Sub-group analysis, children with repeated (longitudinal) measurements⁑ | | Age stratified, children aged >24 months old⁂ |
| --- | --- | --- | --- | --- | --- | --- | --- |
|  | 12-month | 24-month | 12-month | 24-month | 12-month | 24-month | 24-month |
| Any bacterial or protozoan infection | 1.05 (0.96 - 1.15), p=0.29 | 1.00 (0.92 - 1.09), p=0.97 | 0.95 (0.64 - 1.42), p=0.81 | 0.97 (0.79 - 1.18), p=0.73 | 1.02 (0.91 - 1.14), p=0.73 | 0.99 (0.89 - 1.11), p=0.89 | 0.98 (0.91 - 1.05), p=0.57 |
| Any STH infection | 1.16 (0.93 - 1.43), p=0.18 | 0.94 (0.77 - 1.15), p=0.54 | 1.38 (0.35 - 5.44), p=0.65 | 0.48 (0.26 - 0.92), p=0.026 | 1.20 (0.91 - 1.59), p=0.20 | 1.22 (0.85 - 1.75), p=0.27 | 1.04 (0.83 - 1.32), p=0.72 |
| Diarrhea | 1.73 (0.91 - 3.28), p=0.094 | 0.84 (0.46 - 1.51), p=0.55 | 1.66 (0.32 - 8.68), p=0.55 | 1.32 (0.45 - 3.90), p=0.61 | 1.71 (0.79 - 3.71), p=0.17 | 0.68 (0.31 - 1.48), p=0.33 | 0.82 (0.36 - 1.87), p=0.64 |
| Any Bacteria | 1.10 (0.96 - 1.26), p=0.15 | 1.01 (0.88 - 1.16), p=0.87 | 1.23 (0.75 - 2.02), p=0.42 | 0.88 (0.66 - 1.16), p=0.37 | 1.02 (0.86 - 1.20), p=0.85 | 1.02 (0.85 - 1.22), p=0.85 | 0.96 (0.84 - 1.11), p=0.61 |
| *Shigella* | 1.14 (0.94 - 1.38), p=0.18 | 0.97 (0.81 - 1.16), p=0.75 | 0.87 (0.25 - 3.02), p=0.83 | 0.48 (0.28 - 0.84), p=0.009 | 1.09 (0.87 - 1.35), p=0.47 | 0.96 (0.75 - 1.23), p=0.76 | 1.02 (0.85 - 1.23), p=0.82 |
| ETEC | 0.97 (0.70 - 1.35), p=0.86 | 0.83 (0.57 - 1.20), p=0.32 | 0.80 (0.33 - 1.95), p=0.63 | 0.84 (0.47 - 1.49), p=0.55 | 0.86 (0.58 - 1.29), p=0.47 | 0.86 (0.52 - 1.40), p=0.53 | 0.75 (0.47 - 1.20), p=0.23 |
| *Campylobacter* | 1.70 (0.83 - 3.49), p=0.15 | 1.29 (0.63 - 2.64), p=0.49 | 2.67 (0.59 - 12.00), p=0.2 | 1.63 (0.59 - 4.54), p=0.35 | 1.51 (0.60 - 3.76), p=0.38 | 1.52 (0.60 - 3.83), p=0.38 | 0.98 (0.30 - 3.21), p=0.97 |
| *C. difficile* | 2.06 (0.76 - 5.53), p=0.15 | 1.38 (0.45 - 4.20), p=0.57 | 1.42 (0.43 - 4.65), p=0.57 | 1.45 (0.40 - 5.25), p=0.57 | 1.35 (0.23 - 7.78), p=0.74 | 0.23 (0.02 - 2.67), p=0.24 | ..‡ |
| *E. coli* O157 | 0.47 (0.18 - 1.23), p=0.13 | 0.52 (0.17 - 1.59), p=0.25 | 0.00 (0.00 - 0.01), p=0.00 | 0.52 (0.07 - 4.14), p=0.54 | 0.68 (0.22 - 2.07), p=0.50 | 0.58 (0.12 - 2.86), p=0.51 | 0.48 (0.13 - 1.78), p=0.27 |
| STEC | 0.15 (0.03 - 0.71), p=0.017 | 0.24 (0.06 - 1.03), p=0.055 | ..‡ | 0.05 (0.00 - 1.26), p=0.069 | 0.11 (0.01 - 1.32), p=0.082 | 0.58 (0.07 - 5.00), p=0.62 | 1.70 (0.14 - 20.35), p=0.67 |
| *Y. enterocolitica* | ..‡ | ..‡ | ..‡ | ..‡ | ..‡ | ..‡ | ..‡ |
| *V. cholerae* | ..‡ | ..‡ | ..‡ | ..‡ | ..‡ | ..‡ | ..‡ |
| Any Protozoa | 1.05 (0.89 - 1.23), p=0.6 | 0.92 (0.78 - 1.09), p=0.34 | 0.42 (0.14 - 1.26), p=0.12 | 0.86 (0.60 - 1.23), p=0.41 | 1.20 (0.97 - 1.48), p=0.095 | 0.92 (0.73 - 1.16), p=0.49 | 0.94 (0.80 - 1.10), p=0.45 |
| *Giardia* | 1.07 (0.91 - 1.26), p=0.43 | 0.95 (0.80 - 1.12), p=0.51 | 0.46 (0.15 - 1.47), p=0.19 | 0.89 (0.62 - 1.28), p=0.52 | 1.19 (0.96 - 1.47), p=0.11 | 0.92 (0.73 - 1.16), p=0.47 | 0.96 (0.81 - 1.13), p=0.6 |
| *Cryptosporidium* | 0.89 (0.24 - 3.33), p=0.86 | 0.53 (0.13 - 2.17), p=0.38 | 0.33 (0.02 - 6.28), p=0.46 | 0.51 (0.09 - 2.78), p=0.44 | 1.46 (0.21 - 10.18), p=0.7 | 0.59 (0.06 - 5.45), p=0.64 | 0.20 (0.02 - 2.28), p=0.19 |
| *E. histolytica* | ..‡ | ..‡ | ..‡ | ..‡ | ..‡ | ..‡ | ..‡ |
| Any virus | 0.75 (0.44 - 1.28), p=0.29 | 1.03 (0.57 - 1.86), p=0.92 | 0.37 (0.14 - 1.03), p=0.056 | 0.79 (0.35 - 1.78), p=0.57 | 1.09 (0.52 - 2.29), p=0.83 | 0.95 (0.41 - 2.19), p=0.91 | 1.44 (0.61 - 3.38), p=0.41 |
| Norovirus GI/GII | 0.68 (0.36 - 1.28), p=0.23 | 1.10 (0.55 - 2.18), p=0.79 | 0.42 (0.12 - 1.41), p=0.16 | 1.25 (0.47 - 3.29), p=0.66 | 0.86 (0.37 - 2.00), p=0.73 | 0.74 (0.29 - 1.90), p=0.53 | 1.16 (0.45 - 3.04), p=0.76 |
| Adenovirus 40/41 | 1.26 (0.32 - 4.95), p=0.74 | 0.96 (0.18 - 5.20), p=0.96 | 0.85 (0.09 - 8.30), p=0.89 | ..‡ | 3.77 (0.48 - 29.56), p=0.21 | 6.17 (0.51 - 75.19), p=0.15 | 7.51 (0.72 - 77.98), p=0.091 |
| Rotavirus A | ..‡ | ..‡ | ..‡ | ..‡ | ..‡ | ..‡ | ..‡ |
| Coinfection, ≥2 GPP pathogens | 1.10 (0.93 - 1.30), p=0.27 | 0.94 (0.80 - 1.11), p=0.49 | 0.75 (0.33 - 1.71), p=0.49 | 0.83 (0.58 - 1.17), p=0.29 | 1.15 (0.93 - 1.42), p=0.19 | 0.97 (0.78 - 1.21), p=0.81 | 0.93 (0.78 - 1.11), p=0.44 |
| *Trichuris* | 1.05 (0.83 - 1.32), p=0.68 | 0.85 (0.67 - 1.08), p=0.17 | 0.99 (0.23 - 4.27), p=0.98 | 0.24 (0.10 - 0.60), p=0.002 | 1.11 (0.80 - 1.52), p=0.54 | 1.14 (0.76 - 1.70), p=0.54 | 0.99 (0.77 - 1.27), p=0.92 |
| *Ascaris* | 1.38 (0.95 - 1.99), p=0.088 | 0.83 (0.54 - 1.26), p=0.37 | 3.11 (0.30 - 32.54), p=0.34 | 0.65 (0.29 - 1.47), p=0.3 | 1.20 (0.76 - 1.92), p=0.43 | 0.86 (0.42 - 1.75), p=0.68 | 0.86 (0.51 - 1.44), p=0.56 |
| Coinfection, ≥2 STH | 1.21 (0.78 - 1.85), p=0.39 | 0.62 (0.37 - 1.06), p=0.079 | 1.76 (0.15 - 21), p=0.66 | 0.12 (0.01 - 1.06), p=0.057 | 1.01 (0.53 - 1.93), p=0.97 | 0.70 (0.30 - 1.62), p=0.40 | 0.72 (0.40 - 1.29), p=0.27 |

All effect estimates are presented as prevalence ratios (ratio of ratios) with 95% confidence intervals and estimated using generalized estimating equations to fit Poisson regression models with robust standard errors. All models are adjusted for a priori covariates (age, sex, wealth, caregiver education) and age squared to assess the impact of the age squared term on effect estimates. †Results from main analyses examining intervention effects among all enrolled children at 12-month and 24-month visits. Effect estimates compared with 12-month and 24-month results in Table 2. *Results from sub-group analyses which compared children born after the intervention was implemented with children of a similar age at baseline. Effect estimates compared with results in Table 3 (24-month sub-group analysis results) and Supplemental Table 13 (12-month sub-group analysis results). ⁑Results from sub-group analyses including children with repeated measures at baseline and the 12-month phase or baseline and the 24-month phase. Effect estimates compared with results in Supplemental Tables 14 and 15. ⁂ Results from sub-group analysis comparing children aged >2 years old at baseline and 24-month phase. Effect estimates compared with results in Supplemental Table 12. Source files available in Supplemental Table 10 - source data 1 and Supplemental Table 10 - source code 1.

### Supplemental Table 11: Comparison of effect estimates (prevalence ratios) at 12- and 24 month adjusted for *a priori* covariates only and for *a priori* covariates and seasonality.

|  | 12-month prevalence ratio (95% CI) | | 24-month prevalence ratio (95% CI) | |
| --- | --- | --- | --- | --- |
|  | Adjusted (*a priori* only)† | Adjusted + Seasonality* | Adjusted (*a priori* only)† | Adjusted + Seasonality* |
| Any bacterial or protozoan infection | 1.04 (0.94 - 1.15), p=0.41 | 1.05 (0.95 - 1.15), p=0.37 | 0.99 (0.91 - 1.09), p=0.89 | 1.00 (0.91 - 1.10), p=0.95 |
| Any STH infection | 1.11 (0.89 - 1.38), p=0.35 | 1.12 (0.90 - 1.39), p=0.31 | 0.95 (0.77 - 1.17), p=0.62 | 0.94 (0.76 - 1.15), p=0.54 |
| Diarrhea | 1.69 (0.89 - 3.21), p=0.11 | 1.67 (0.88 - 3.17), p=0.12 | 0.84 (0.47 - 1.51), p=0.56 | 0.81 (0.44 - 1.46), p=0.48 |
| Any Bacteria | 1.09 (0.95 - 1.26), p=0.20 | 1.10 (0.96 - 1.26), p=0.18 | 1.00 (0.87 - 1.15), p=0.95 | 1.03 (0.89 - 1.18), p=0.71 |
| *Shigella* | 1.12 (0.92 - 1.38), p=0.27 | 1.12 (0.91 - 1.37), p=0.28 | 0.95 (0.79 - 1.16), p=0.64 | 0.97 (0.80 - 1.17), p=0.72 |
| ETEC | 0.96 (0.69 - 1.33), p=0.81 | 0.98 (0.70 - 1.35), p=0.89 | 0.83 (0.57 - 1.19), p=0.31 | 0.88 (0.61 - 1.26), p=0.47 |
| *Campylobacter* | 1.68 (0.82 - 3.45), p=0.16 | 1.72 (0.84 - 3.49), p=0.14 | 1.28 (0.62 - 2.62), p=0.5 | 1.33 (0.65 - 2.71), p=0.43 |
| *C. difficile* | 2.09 (0.77 - 5.64), p=0.15 | 2.17 (0.81 - 5.86), p=0.13 | 1.41 (0.46 - 4.30), p=0.54 | 1.44 (0.48 - 4.37), p=0.52 |
| *E. coli* O157 | 0.46 (0.18 - 1.21), p=0.12 | 0.48 (0.18 - 1.26), p=0.14 | 0.52 (0.17 - 1.59), p=0.25 | 0.57 (0.19 - 1.74), p=0.32 |
| STEC | 0.15 (0.03 - 0.70), p=0.016 | 0.15 (0.03 - 0.74), p=0.019 | 0.24 (0.05 - 1.01), p=0.052 | 0.25 (0.06 - 1.06), p=0.061 |
| *Y. enterocolitica* | ..‡ | ..‡ | ..‡ | ..‡ |
| *V. cholerae* | ..‡ | ..‡ | ..‡ | ..‡ |
| Any Protozoa | 1.03 (0.86 - 1.22), p=0.76 | 1.03 (0.87 - 1.23), p=0.72 | 0.91 (0.76 - 1.09), p=0.29 | 0.91 (0.76 - 1.09), p=0.31 |
| *Giardia* | 1.05 (0.88 - 1.25), p=0.58 | 1.06 (0.88 - 1.26), p=0.54 | 0.93 (0.78 - 1.11), p=0.43 | 0.93 (0.78 - 1.12), p=0.45 |
| *Cryptosporidium* | 0.89 (0.24 - 3.31), p=0.86 | 0.83 (0.22 - 3.11), p=0.78 | 0.53 (0.13 - 2.14), p=0.37 | 0.46 (0.12 - 1.73), p=0.25 |
| *E. histolytica* | ..‡ | ..‡ | ..‡ | ..‡ |
| Any virus | 0.75 (0.44 - 1.27), p=0.29 | 0.74 (0.43 - 1.26), p=0.26 | 1.03 (0.57 - 1.86), p=0.92 | 0.97 (0.54 - 1.75), p=0.91 |
| Norovirus GI/GII | 0.68 (0.36 - 1.27), p=0.23 | 0.67 (0.35 - 1.27), p=0.22 | 1.10 (0.55 - 2.18), p=0.79 | 1.04 (0.53 - 2.07), p=0.90 |
| Adenovirus 40/41 | 1.24 (0.32 - 4.83), p=0.76 | 1.29 (0.33 - 5.13), p=0.71 | 0.97 (0.18 - 5.19), p=0.97 | 1.01 (0.19 - 5.30), p=0.99 |
| Rotavirus | ..‡ | ..‡ | ..‡ | ..‡ |
| Coinfection, ≥2 GPP pathogens | 1.08 (0.91 - 1.29), p=0.37 | 1.09 (0.91 - 1.30), p=0.35 | 0.93 (0.79 - 1.10), p=0.41 | 0.94 (0.79 - 1.12), p=0.49 |
| *Trichuris* | 1.01 (0.79 - 1.28), p=0.96 | 1.02 (0.81 - 1.30), p=0.86 | 0.86 (0.67 - 1.10), p=0.22 | 0.85 (0.67 - 1.09), p=0.21 |
| *Ascaris* | 1.33 (0.92 - 1.93), p=0.13 | 1.35 (0.93 - 1.95), p=0.11 | 0.83 (0.54 - 1.27), p=0.39 | 0.81 (0.53 - 1.25), p=0.34 |
| Coinfection, ≥2 STH | 1.17 (0.76 - 1.79), p=0.49 | 1.20 (0.78 - 1.83), p=0.40 | 0.63 (0.37 - 1.07), p=0.084 | 0.62 (0.36 - 1.06), p=0.079 |

All effect estimates are presented as prevalence ratios (ratio of ratios) with 95% confidence intervals and estimated using generalized estimating equations to fit Poisson regression models with robust standard errors. †Models are adjusted for *a priori* covariates age, sex, caregiver’s education, and wealth and presented for comparison with seasonality-adjusted models. *Models are adjusted for *a priori* covariates and seasonality using sine/cosine terms based on the date of sample (or survey) collection. Source files available in Supplemental Table 11 - source data 1 and Supplemental Table 11 - source code 1.

### Supplemental Table 12: Effect of the intervention on enteric infection and diarrhea in children >2 years old after 24 months

|  | **Prevalence** | | **Prevalence ratio (95% CI), p-value** | |
| --- | --- | --- | --- | --- |
|  | **Baseline, aged >2 years** | **24-month, aged >2 years** | **unadjusted** | **adjusted**† |
| Any bacterial or protozoan infection‡ |  |  |  |  |
| Control | 155/164 (95%) | 315/340 (93%) | .. | .. |
| Intervention | 149/160 (93%) | 312/344 (91%) | 0.99 (0.93 - 1.07), p=0.86 | 0.98 (0.91 - 1.05), p=0.60 |
| Any STH infection‡ |  |  |  |  |
| Control | 103/155 (66%) | 113/175 (65%) | .. | .. |
| Intervention | 86/146 (59%) | 121/208 (58%) | 1.03 (0.82 - 1.30), p=0.79 | 1.05 (0.83 - 1.32), p=0.69 |
| Diarrhea‡ |  |  |  |  |
| Control | 21/243 (8.6%) | 33/273 (12%) | .. | .. |
| Intervention | 16/210 (7.6%) | 31/303 (10%) | 0.96 (0.45 - 2.07), p=0.93 | 0.82 (0.36 - 1.86), p=0.63 |
| Any Bacteria |  |  |  |  |
| Control | 129/164 (79%) | 267/340 (79%) | .. | .. |
| Intervention | 125/160 (78%) | 266/344 (77%) | 1.00 (0.87 - 1.15), p=0.98 | 0.97 (0.84 - 1.11), p=0.64 |
| *Shigella* |  |  |  |  |
| Control | 112/164 (68%) | 227/340 (67%) | .. | .. |
| Intervention | 103/160 (64%) | 223/344 (65%) | 1.05 (0.87 - 1.26), p=0.63 | 1.03 (0.85 - 1.24), p=0.79 |
| ETEC |  |  |  |  |
| Control | 46/164 (28%) | 93/340 (27%) | .. | .. |
| Intervention | 52/160 (33%) | 100/344 (29%) | 0.88 (0.56 - 1.38), p=0.58 | 0.74 (0.46 - 1.20), p=0.22 |
| *Campylobacter* |  |  |  |  |
| Control | 12/164 (7.3%) | 33/340 (9.7%) | .. | .. |
| Intervention | 7/160 (4.4%) | 20/344 (5.8%) | 0.97 (0.33 - 2.90), p=0.96 | 1.00 (0.30 - 3.28), p=0.99 |
| *C. difficile* |  |  |  |  |
| Control | 2/164 (1.2%) | 6/340 (1.8%) | .. | .. |
| Intervention | 0/160 (0.0%) | 4/344 (1.2%) | ..‡ | ..‡ |
| *E. coli* O157 |  |  |  |  |
| Control | 6/164 (3.7%) | 21/340 (6.2%) | .. | .. |
| Intervention | 9/160 (5.6%) | 13/344 (3.8%) | 0.39 (0.11 - 1.40), p=0.15 | 0.47 (0.13 - 1.78), p=0.27 |
| STEC |  |  |  |  |
| Control | 2/164 (1.2%) | 15/340 (4.4%) | .. | .. |
| Intervention | 1/160 (0.63%) | 13/344 (3.8%) | 1.54 (0.12 - 19.19), p=0.74 | 1.73 (0.14 - 20.75), p=0.67 |
| *Y. enterocolitica* |  |  |  |  |
| Control | 0/164 (0.0%) | 0/340 (0.0%) | .. | .. |
| Intervention | 0/160 (0.0%) | 1/344 (0.29%) | ..‡ | ..‡ |
| *V. cholerae* |  |  |  |  |
| Control | 0/164 (0.0%) | 0/340 (0.0%) | .. | .. |
| Intervention | 0/160 (0.0%) | 0/344 (0.0%) | ..‡ | ..‡ |
| Any Protozoa |  |  |  |  |
| Control | 123/164 (75%) | 250/340 (74%) | .. | .. |
| Intervention | 121/160 (76%) | 245/344 (71%) | 0.96 (0.82 - 1.13), p=0.66 | 0.94 (0.80 - 1.11), p=0.47 |
| *Giardia* |  |  |  |  |
| Control | 122/164 (74%) | 244/340 (72%) | .. | .. |
| Intervention | 118/160 (74%) | 240/344 (70%) | 0.99 (0.84 - 1.16), p=0.86 | 0.96 (0.81 - 1.13), p=0.62 |
| *Cryptosporidium* |  |  |  |  |
| Control | 1/164 (0.61%) | 9/340 (2.6%) | .. | .. |
| Intervention | 4/160 (2.5%) | 8/344 (2.3%) | 0.20 (0.02 - 2.27), p=0.19 | 0.21 (0.02 - 2.46), p=0.21 |
| *E. histolytica* |  |  |  |  |
| Control | 0/164 (0.0%) | 2/340 (0.59%) | .. | .. |
| Intervention | 3/160 (1.9%) | 10/344 (2.9%) | ..‡ | ..‡ |
| Any virus |  |  |  |  |
| Control | 19/164 (12%) | 39/340 (11%) | .. | .. |
| Intervention | 16/160 (10%) | 43/344 (13%) | 1.24 (0.55 - 2.78), p=0.6 | 1.44 (0.61 - 3.38), p=0.41 |
| Norovirus GI/GII |  |  |  |  |
| Control | 12/164 (7.3%) | 34/340 (10%) | .. | .. |
| Intervention | 13/160 (8.1%) | 37/344 (11%) | 0.96 (0.39 - 2.34), p=0.92 | 1.17 (0.45 - 3.03), p=0.75 |
| Adenovirus 40/41 |  |  |  |  |
| Control | 6/164 (3.7%) | 2/340 (0.59%) | .. | .. |
| Intervention | 2/160 (1.3%) | 6/344 (1.7%) | 11 (0.97 – 119), p=0.053 | 7.5 (0.72 – 79), p=0.92 |
| Rotavirus A |  |  |  |  |
| Control | 1/164 (0.61%) | 3/340 (0.88%) | .. | .. |
| Intervention | 1/160 (0.63%) | 1/344 (0.29%) | ..‡ | ..‡ |
| Coinfection, ≥2 GPP pathogens |  |  |  |  |
| Control | 114/164 (70%) | 243/340 (71%) | .. | .. |
| Intervention | 111/160 (69%) | 236/344 (69%) | 0.97 (0.82 - 1.15), p=0.71 | 0.93 (0.78 - 1.12), p=0.45 |
| *Trichuris* |  |  |  |  |
| Control | 91/155 (59%) | 102/175 (58%) | .. | .. |
| Intervention | 76/146 (52%) | 110/208 (53%) | 1.04 (0.81 - 1.33), p=0.78 | 0.99 (0.77 - 1.27), p=0.96 |
| *Ascaris* |  |  |  |  |
| Control | 50/155 (32%) | 61/175 (35%) | .. | .. |
| Intervention | 39/146 (27%) | 47/208 (23%) | 0.78 (0.47 - 1.29), p=0.33 | 0.86 (0.51 - 1.44), p=0.57 |
| Coinfection, ≥2 STH |  |  |  |  |
| Control | 38/155 (25%) | 50/175 (29%) | .. | .. |
| Intervention | 29/146 (20%) | 36/208 (17%) | 0.74 (0.42 - 1.28), p=0.28 | 0.72 (0.41 – 1.29), p=0.27 |

Analysis includes children <2 year old at baseline or the 24-month visit. Prevalence results are presented as (n/N (%)). All effect estimates are presented as prevalence ratios (ratio of ratios) with 95% confidence intervals and estimated using generalized estimating equations to fit Poisson regression models with robust standard errors. †Pathogen outcomes adjusted for child age and sex, caregiver’s education, and household wealth index, reported diarrhea also adjusted for baseline presence of a drop-hole cover and reported use of a tap on compound grounds as primary drinking water source. ‡ Models did not converge due to sparse data. Source files available in Supplemental Table 12 - source data 1 and Supplemental Table 12 - source code 1

Supplemental Table 13: Effect of intervention on enteric infection and reported diarrhea in children born into study sites post implementation (post-baseline) and before 12-month visit compared with children of a similar age at baseline (<1 year old).

|  | **Prevalence** | | **Prevalence ratio** | |
| --- | --- | --- | --- | --- |
|  | **Baseline, children <1 year old** | **12-month, children born-in & <1 year old** | **unadjusted** | **adjusted†** |
| Any bacterial or protozoan infection |  |  |  |  |
| Control | 57/109 (52%) | 31/48 (65%) | .. | .. |
| Intervention | 51/99 (52%) | 32/55 (58%) | 0.89 (0.60 - 1.33), p=0.58 | 0.97 (0.65 - 1.45), p=0.90 |
| Any STH infection |  |  |  |  |
| Control | 17/93 (18%) | 3/25 (12%) | .. | .. |
| Intervention | 13/92 (14%) | 4/32 (13%) | 1.31 (0.32 - 5.42), p=0.71 | 1.38 (0.35 - 5.45), p=0.65 |
| Diarrhea |  |  |  |  |
| Control | 19/138 (14%) | 6/50 (12%) | .. | .. |
| Intervention | 18/120 (15%) | 13/69 (19%) | 1.38 (0.47 - 4.01), p=0.56 | 1.80 (0.35 - 9.31), p=0.48 |
| Any Bacteria |  |  |  |  |
| Control | 53/109 (49%) | 24/48 (50%) | .. | .. |
| Intervention | 41/99 (41%) | 29/55 (53%) | 1.22 (0.75 - 1.98), p=0.43 | 1.28 (0.78 - 2.10), p=0.33 |
| *Shigella* |  |  |  |  |
| Control | 10/109 (9.2%) | 9/48 (19%) | .. | .. |
| Intervention | 9/99 (9.1%) | 9/55 (16%) | 0.87 (0.26 - 2.91), p=0.82 | 0.85 (0.26 - 2.81), p=0.79 |
| ETEC |  |  |  |  |
| Control | 25/109 (23%) | 12/48 (25%) | .. | .. |
| Intervention | 22/99 (22%) | 11/55 (20%) | 0.82 (0.34 - 1.99), p=0.66 | 0.80 (0.33 - 1.92), p=0.62 |
| *Campylobacter* |  |  |  |  |
| Control | 14/109 (13%) | 4/48 (8.3%) | .. | .. |
| Intervention | 8/99 (8.1%) | 5/55 (9.1%) | 1.76 (0.38 - 8.09), p=0.47 | 2.68 (0.59 - 12.2), p=0.20 |
| *C. difficile* |  |  |  |  |
| Control | 13/109 (12%) | 7/48 (15%) | .. | .. |
| Intervention | 10/99 (10%) | 9/55 (16%) | 1.37 (0.42 - 4.45), p=0.60 | 1.49 (0.46 - 4.89), p=0.51 |
| E. coli O157 |  |  |  |  |
| Control | 4/109 (3.7%) | 1/48 (2.1%) | .. | .. |
| Intervention | 2/99 (2%) | 0/55 (0.0%) | 0.01 (0.00 - 0.19), p=0.001 | ..‡ |
| STEC |  |  |  |  |
| Control | 0/109 (0.0%) | 0/48 (0.0%) | .. | .. |
| Intervention | 3/99 (3%) | 1/55 (1.8%) | ..‡ | ..‡ |
| *Y. enterocolitica* |  |  |  |  |
| Control | 0/109 (0.0%) | 0/48 (0.0%) | .. | .. |
| Intervention | 0/99 (0.0%) | 0/55 (0.0%) | ..‡ | ..‡ |
| *V. cholerae* |  |  |  |  |
| Control | 0/109 (0.0%) | 0/48 (0.0%) | .. | .. |
| Intervention | 0/99 (0.0%) | 0/55 (0.0%) | ..‡ | ..‡ |
| Any Protozoa |  |  |  |  |
| Control | 14/109 (13%) | 15/48 (31%) | .. | .. |
| Intervention | 22/99 (22%) | 9/55 (16%) | 0.35 (0.12 - 1.02), p=0.055 | 0.40 (0.13 – 1.20), p=0.10 |
| *Giardia* |  |  |  |  |
| Control | 12/109 (11%) | 13/48 (27%) | .. | .. |
| Intervention | 16/99 (16%) | 8/55 (15%) | 0.41 (0.13 - 1.24), p=0.11 | 0.44 (0.14 – 1.40), p=0.17 |
| *Cryptosporidium* |  |  |  |  |
| Control | 2/109 (1.8%) | 2/48 (4.2%) | .. | .. |
| Intervention | 8/99 (8.1%) | 2/55 (3.6%) | 0.25 (0.02 - 3.70), p=0.31 | 0.40 (0.02 – 7.9), p=0.55 |
| *E. histolytica* |  |  |  |  |
| Control | 0/109 (0.0%) | 1/48 (2.1%) | .. | .. |
| Intervention | 1/99 (1%) | 0/55 (0.0%) | ..‡ | ..‡ |
| Any virus |  |  |  |  |
| Control | 15/109 (14%) | 12/48 (25%) | .. | .. |
| Intervention | 21/99 (21%) | 7/55 (13%) | 0.33 (0.12 - 0.92), p=0.033 | 0.37 (0.14 – 1.03), p=0.056 |
| Norovirus GI/GII |  |  |  |  |
| Control | 12/109 (11%) | 9/48 (19%) | .. | .. |
| Intervention | 15/99 (15%) | 6/55 (11%) | 0.43 (0.13 - 1.40), p=0.16 | 0.44 (0.13 – 1.47), p=0.18 |
| Adenovirus 40/41 |  |  |  |  |
| Control | 4/109 (3.7%) | 4/48 (8.3%) | .. | .. |
| Intervention | 3/99 (3%) | 2/55 (3.6%) | 0.56 (0.06 - 5.05), p=0.61 | 0.91 (0.09 - 9.49), p=0.94 |
| Rotavirus A |  |  |  |  |
| Control | 0/109 (0.0%) | 0/48 (0.0%) | .. | .. |
| Intervention | 3/99 (3%) | 0/55 (0.0%) | ..‡ | ..‡ |
| Coinfection, ≥2 GPP pathogens |  |  |  |  |
| Control | 23/109 (21%) | 16/48 (33%) | .. | .. |
| Intervention | 25/99 (25%) | 15/55 (27%) | 0.73 (0.31 - 1.71), p=0.47 | 0.74 (0.33 – 1.69), p=0.48 |
| *Trichuris* |  |  |  |  |
| Control | 10/93 (11%) | 3/25 (12%) | .. | .. |
| Intervention | 10/92 (11%) | 4/32 (13%) | 1.04 (0.21 - 5.01), p=0.96 | 0.98 (0.23 - 4.29), p=0.98 |
| *Ascaris* |  |  |  |  |
| Control | 12/93 (13%) | 1/25 (4%) | .. | .. |
| Intervention | 9/92 (9.8%) | 3/32 (9.4%) | 2.87 (0.30 - 27.85), p=0.36 | 3.10 (0.30 – 32.5), p=0.35 |
| Coinfection, ≥2 STH |  |  |  |  |
| Control | 5/93 (5.4%) | 1/25 (4%) | .. | .. |
| Intervention | 6/92 (6.5%) | 3/32 (9.4%) | 1.90 (0.16 - 22.73), p=0.61 | 1.76 (0.15 – 21.0), p=0.66 |

Analysis includes children <1 year old at baseline and children born into the study after baseline and <1 year old at the time of the 12-month visit. Prevalence results are presented as (n/N (%)). All effect estimates are presented as prevalence ratios (ratio of ratios) with 95% confidence intervals and estimated using generalized estimating equations to fit Poisson regression models with robust standard errors. †Pathogen outcomes adjusted for child age and sex, caregiver’s education, and household wealth index, reported diarrhea also adjusted for baseline presence of a drop-hole cover and reported use of a tap on compound grounds as primary drinking water source. ‡ Models did not converge due to sparse data. Source files available in Supplemental Table 13 - source data 1 and Supplemental Table 13 - source code

### Supplemental Table 14: Effect of the intervention on children with repeated observations at baseline and 12-month visit.

|  | Prevalence | | Prevalence ratio | |
| --- | --- | --- | --- | --- |
|  | Baseline | 12-month | unadjusted | adjusted† |
| Any bacterial or protozoan infection |  |  |  |  |
| Control | 161/207 (78%) | 187/207 (90%) | .. | .. |
| Intervention | 174/228 (76%) | 207/228 (91%) | 1.02 (0.91 - 1.16), p=0.70 | 1.01 (0.90 - 1.14), p=0.84 |
| Any STH infection |  |  |  |  |
| Control | 67/132 (51%) | 80/132 (61%) | .. | .. |
| Intervention | 63/154 (41%) | 91/154 (59%) | 1.22 (0.92 - 1.61), p=0.17 | 1.16 (0.87 - 1.55), p=0.31 |
| Diarrhea |  |  |  |  |
| Control | 36/277 (13%) | 17/277 (6.1%) | .. | .. |
| Intervention | 42/279 (15%) | 34/279 (12%) | 1.71 (0.78 - 3.77), p=0.18 | 1.71 (0.79 - 3.70), p=0.17 |
| Any Bacteria |  |  |  |  |
| Control | 141/207 (68%) | 165/207 (80%) | .. | .. |
| Intervention | 142/228 (62%) | 170/228 (75%) | 1.02 (0.86 - 1.22), p=0.8 | 1.01 (0.85 - 1.20), p=0.92 |
| *Shigella* |  |  |  |  |
| Control | 89/207 (43%) | 128/207 (62%) |  |  |
| Intervention | 90/228 (39%) | 142/228 (62%) | 1.10 (0.86 - 1.39), p=0.45 | 1.08 (0.85 - 1.37), p=0.54 |
| *ETEC* |  |  |  |  |
| Control | 63/207 (30%) | 83/207 (40%) |  |  |
| Intervention | 71/228 (31%) | 79/228 (35%) | 0.84 (0.56 - 1.27), p=0.41 | 0.85 (0.57 - 1.28), p=0.44 |
| *Campylobacter* |  |  |  |  |
| Control | 20/207 (9.7%) | 18/207 (8.7%) |  |  |
| Intervention | 13/228 (5.7%) | 18/228 (7.9%) | 1.54 (0.62 - 3.80), p=0.35 | 1.49 (0.60 - 3.71), p=0.39 |
| *C. difficile* |  |  |  |  |
| Control | 15/207 (7.3%) | 4/207 (1.9%) |  |  |
| Intervention | 8/228 (3.5%) | 3/228 (1.3%) | 1.39 (0.24 - 8.00), p=0.71 | 1.45 (0.25 - 8.52), p=0.68 |
| *E. coli* O157 |  |  |  |  |
| Control | 9/207 (4.3%) | 15/207 (7.3%) | .. | .. |
| Intervention | 9/228 (4.0%) | 10/228 (4.4%) | 0.67 (0.22 - 2.03), p=0.48 | 0.68 (0.22 - 2.06), p=0.49 |
| STEC |  |  |  |  |
| Control | 1/207 (0.48%) | 6/207 (2.9%) | .. | .. |
| Intervention | 6/228 (2.6%) | 4/227 (1.8%) | 0.11 (0.01 - 1.31), p=0.081 | 0.11 (0.01 - 1.32), p=0.082 |
| *Y. enterocolitica* |  |  |  |  |
| Control | 0/207 (0.0%) | 0/207 (0.0%) | .. | .. |
| Intervention | 1/228 (0.44%) | 0/227 (0.0%) | ..‡ | ..‡ |
| *V. cholerae* |  |  |  |  |
| Control | 0/207 (0.0%) | 0/207 (0.0%) | .. | .. |
| Intervention | 0/228 (0.0%) | 0/227 (0.0%) | ..‡ | ..‡ |
| Any Protozoa |  |  |  |  |
| Control | 109/207 (53%) | 130/207 (63%) | .. | .. |
| Intervention | 117/228 (51%) | 166/228 (73%) | 1.19 (0.95 - 1.48), p=0.13 | 1.18 (0.94 - 1.47), p=0.15 |
| *Giardia* |  |  |  |  |
| Control | 106/207 (51%) | 130/207 (63%) |  |  |
| Intervention | 113/228 (50%) | 164/228 (72%) | 1.18 (0.94 - 1.48), p=0.15 | 1.17 (0.93 - 1.47), p=0.17 |
| *Cryptosporidium* |  |  |  |  |
| Control | 6/207 (2.9%) | 2/207 (0.97%) | .. | .. |
| Intervention | 10/228 (4.4%) | 5/227 (2.2%) | 1.44 (0.21 - 9.82), p=0.71 | 1.45 (0.22 - 9.71), p=0.7 |
| *E. histolytica* |  |  |  |  |
| Control | 0/207 (0.0%) | 0/207 (0.0) | .. | .. |
| Intervention | 2/228 (0.88%) | 7/228 (3.1%) | ..‡ | ..‡ |
| Any virus |  |  |  |  |
| Control | 27/207 (13%) | 20/207 (9.7%) | .. | .. |
| Intervention | 31/228 (14%) | 25/228 (11%) | 1.05 (0.50 - 2.22), p=0.89 | 1.08 (0.51 - 2.26), p=0.84 |
| Norovirus GI/GII |  |  |  |  |
| Control | 20/207 (9.7%) | 19/207 (9.2%) |  |  |
| Intervention | 23/228 (11%) | 19/228 (8.3%) | 0.83 (0.36 - 1.94), p=0.67 | 0.86 (0.37 - 1.99), p=0.72 |
| Adenovirus 40/41 |  |  |  |  |
| Control | 7/207 (3.4%) | 2/207 (0.97%) | .. | .. |
| Intervention | 6/228 (2.6%) | 6/228 (2.6%) | 3.56 (0.46 - 27.24), p=0.22 | 3.59 (0.46 - 27.91), p=0.22 |
| Rotavirus A |  |  |  |  |
| Control | 1/207 (0.48%) | 1/207 (0.48%) | .. | .. |
| Intervention | 4/228 (1.8%) | 1/228 (0.44%) | ..‡ | ..‡ |
| Coinfection, ≥2 GPP pathogens |  |  |  |  |
| Control | 114/207 (55%) | 135/207 (65%) | .. | .. |
| Intervention | 115/228 (50%) | 156/228 (68%) | 1.15 (0.92 - 1.43), p=0.23 | 1.14 (0.91 - 1.42), p=0.25 |
| *Trichuris* |  |  |  |  |
| Control | 49/132 (37%) | 64/132 (48%) | .. | .. |
| Intervention | 53/154 (34%) | 77/154 (50%) | 1.12 (0.81 - 1.54), p=0.50 | 1.06 (0.76 - 1.48), p=0.72 |
| *Ascaris* |  |  |  |  |
| Control | 40/132 (30%) | 46/132 (35%) |  |  |
| Intervention | 35/154 (23%) | 49/154 (32%) | 1.22 (0.77 - 1.93), p=0.4 | 1.17 (0.73 - 1.86), p=0.51 |
| Coinfection, ≥2 STH |  |  |  |  |
| Control | 22/132 (17%) | 30/132 (23%) | .. | .. |
| Intervention | 25/154 (16%) | 35/154 (23%) | 1.03 (0.55 - 1.93), p=0.94 | 0.97 (0.51 - 1.85), p=0.93 |

Analysis includes children with complete observations at baseline and 12-month visits. Prevalence results are presented as (n/N (%)). All effect estimates are presented as prevalence ratios (ratio of ratios) with 95% confidence intervals and estimated using generalized estimating equations to fit Poisson regression models with robust standard errors. †Pathogen outcomes adjusted for child age and sex, caregiver’s education, and household wealth index, reported diarrhea also adjusted for baseline presence of a drop-hole cover and reported use of a tap on compound grounds as primary drinking water source. ‡ Models would not converge due to sparse data. Source files available in Supplemental Table 14 - source data 1 and Supplemental Table 14 - source code

### Supplemental Table 15: Effect of the intervention on children with repeated observations at baseline and 24-month visit.

|  | **Prevalence** | | **Prevalence ratio** | |
| --- | --- | --- | --- | --- |
|  | **Baseline** | **24-month** | **unadjusted** | **adjusted†** |
| Any bacterial or protozoan infection |  |  |  |  |
| Control | 131/166 (79%) | 155/166 (93%) | .. | .. |
| Intervention | 151/192 (79%) | 175/192 (91%) | 0.98 (0.87 - 1.10), p=0.73 | 0.98 (0.87 - 1.10), p=0.70 |
| Any STH infection |  |  |  |  |
| Control | 48/95 (51%) | 65/95 (68%) | .. | .. |
| Intervention | 38/106 (36%) | 62/106 (58%) | 1.20 (0.84 - 1.70), p=0.31 | 1.25 (0.87 - 1.78), p=0.23 |
| Diarrhea |  |  |  |  |
| Control | 25/196 (13%) | 20/196 (10%) | .. | .. |
| Intervention | 34/221 (15%) | 20/221 (9.1%) | 0.72 (0.33 - 1.58), p=0.41 | 0.69 (0.31 - 1.50), p=0.35 |
| Any Bacteria |  |  |  |  |
| Control | 109/166 (66%) | 138/166 (83%) | .. | .. |
| Intervention | 120/192 (63%) | 153/192 (80%) | 1.00 (0.84 - 1.21), p=0.96 | 1.01 (0.83 - 1.21), p=0.96 |
| *Shigella* |  |  |  |  |
| Control | 66/166 (40%) | 121/166 (73%) |  |  |
| Intervention | 79/192 (41%) | 136/192 (71%) | 0.93 (0.71 - 1.22), p=0.60 | 0.93 (0.71 - 1.22), p=0.60 |
| *ETEC* |  |  |  |  |
| Control | 47/166 (28%) | 47/166 (28%) |  |  |
| Intervention | 58/192 (30%) | 52/192 (27%) | 0.90 (0.55 - 1.46), p=0.66 | 0.85 (0.52 - 1.39), p=0.52 |
| *Campylobacter* |  |  |  |  |
| Control | 16/166 (9.6%) | 12/166 (7.2%) |  |  |
| Intervention | 13/192 (6.8%) | 14/192 (7.3%) | 1.44 (0.56 - 3.72), p=0.45 | 1.52 (0.60 - 3.83), p=0.37 |
| *C. difficile* |  |  |  |  |
| Control | 9/166 (5.4%) | 4/166 (2.4%) | .. | .. |
| Intervention | 8/192 (4.2%) | 1/192 (0.52%) | 0.28 (0.03 - 2.95), p=0.29 | 0.26 (0.03 - 2.59), p=0.25 |
| *E. coli* O157 |  |  |  |  |
| Control | 7/166 (4.2%) | 9/166 (5.4%) | .. | .. |
| Intervention | 9/192 (4.7%) | 8/192 (4.2%) | 0.69 (0.14 - 3.40), p=0.65 | 0.59 (0.12 - 2.93), p=0.52 |
| STEC |  |  |  |  |
| Control | 2/166 (1.2%) | 7/166 (4.2%) | .. | .. |
| Intervention | 3/192 (1.6%) | 7/192 (3.6%) | 0.66 (0.07 - 6.20), p=0.72 | 0.58 (0.07 - 4.89), p=0.61 |
| *Y. enterocolitica* |  |  |  |  |
| Control | 0/166 (0.0%) | 0/166 (0.0%) | .. | .. |
| Intervention | 0/192 (0.0%) | 1/192 (0.52%) | ..‡ | ..‡ |
| *V. cholerae* |  |  |  |  |
| Control | 0/166 (0.0%) | 0/166 (0.0%) | .. | .. |
| Intervention | 0/192 (0.0%) | 0/192 (0.0%) | ..‡ | ..‡ |
| Any Protozoa |  |  |  |  |
| Control | 89/166 (54%) | 121/166 (73%) | .. | .. |
| Intervention | 109/192 (57%) | 138/192 (72%) | 0.93 (0.73 - 1.19), p=0.56 | 0.90 (0.69 - 1.15), p=0.39 |
| *Giardia* |  |  |  |  |
| Control | 86/166 (52%) | 120/166 (72%) |  |  |
| Intervention | 104/192 (54%) | 135/192 (70%) | 0.93 (0.73 - 1.18), p=0.55 | 0.89 (0.69 - 1.15), p=0.38 |
| *Cryptosporidium* |  |  |  |  |
| Control | 5/166 (3%) | 3/166 (1.8%) | .. | .. |
| Intervention | 11/192 (5.7%) | 4/192 (2.1%) | 0.57 (0.06 - 5.38), p=0.62 | 0.55 (0.06 - 4.93), p=0.59 |
| *E. histolytica* |  |  |  |  |
| Control | 0/166 (0.0%) | 0/166 (0.0%) | .. | .. |
| Intervention | 2/192 (1%) | 8/192 (4.2%) | ..‡ | ..‡ |
| Any virus |  |  |  |  |
| Control | 21/166 (13%) | 18/166 (11%) | .. | .. |
| Intervention | 30/192 (16%) | 22/192 (11%) | 0.86 (0.37 - 1.97), p=0.72 | 0.95 (0.41 - 2.19), p=0.91 |
| Norovirus GI/GII |  |  |  |  |
| Control | 15/166 (9%) | 15/166 (9%) | .. | .. |
| Intervention | 26/192 (14%) | 17/192 (8.8%) | 0.65 (0.25 - 1.69), p=0.38 | 0.74 (0.28 - 1.90), p=0.53 |
| Adenovirus 40/41 |  |  |  |  |
| Control | 6/166 (3.6%) | 1/166 (0.6%) |  |  |
| Intervention | 5/192 (2.6%) | 5/192 (2.6%) | 6.12 (0.48 - 78.34), p=0.16 | 6.01 (0.49 - 73.94), p=0.16 |
| Rotavirus A |  |  |  |  |
| Control | 1/166 (0.6%) | 2/166 (1.2%) | .. | .. |
| Intervention | 1/192 (0.52%) | 1/192 (0.52%) | ..‡ | ..‡ |
| Coinfection, ≥2 GPP pathogens |  |  |  |  |
| Control | 89/166 (54%) | 120/166 (72%) | .. | .. |
| Intervention | 102/192 (53%) | 132/192 (69%) | 0.96 (0.77 - 1.19), p=0.69 | 0.95 (0.76 - 1.19), p=0.67 |
| *Trichuris* |  |  |  |  |
| Control | 39/95 (41%) | 62/95 (65%) | .. | .. |
| Intervention | 32/106 (30%) | 57/106 (54%) | 1.11 (0.74 - 1.67), p=0.60 | 1.16 (0.77 - 1.75), p=0.47 |
| *Ascaris* |  |  |  |  |
| Control | 27/95 (28%) | 34/95 (36%) |  |  |
| Intervention | 19/106 (18%) | 21/106 (20%) | 0.88 (0.43 - 1.79), p=0.72 | 0.89 (0.44 - 1.79), p=0.74 |
| Coinfection, ≥2 STH |  |  |  |  |
| Control | 18/95 (19%) | 31/95 (33%) | .. | .. |
| Intervention | 13/106 (12%) | 16/106 (15%) | 0.71 (0.30 - 1.70), p=0.44 | 0.72 (0.31 - 1.69), p=0.46 |

Analysis includes children with complete observations at baseline and 24-month visits. Prevalence results are presented as (n/N (%)). All effect estimates are presented as prevalence ratios (ratio of ratios) with 95% confidence intervals and estimated using generalized estimating equations to fit Poisson regression models with robust standard errors. †Pathogen outcomes adjusted for child age and sex, caregiver’s education, and household wealth index, reported diarrhea also adjusted for baseline presence of a drop-hole cover and reported use of a tap on compound grounds as primary drinking water source. ‡ Models would not converge due to sparse data. Source files available in Supplemental Table 15 - source data 1 and Supplemental Table 15 - source code 1.

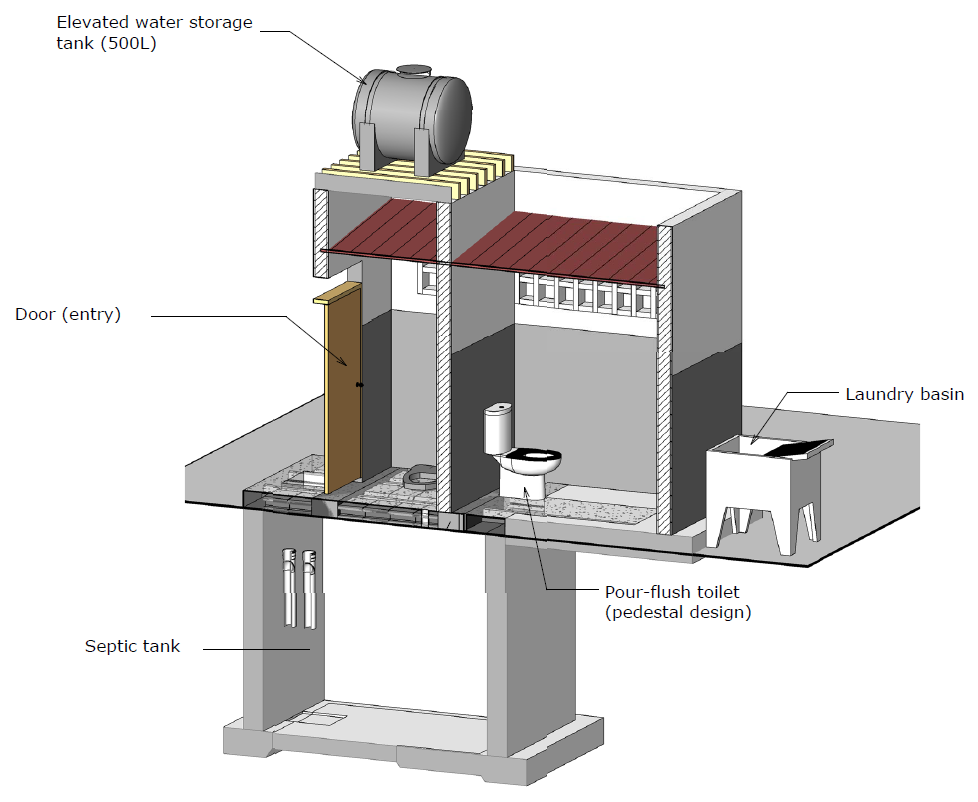

Supplemental Figure 5a: Schematic of communal sanitation block design from the NGO (Water and Sanitation for the Urban Poor). Pictured: 2 latrine stalls, 2 pour-flush toilets, septic tank, elevated water storage tank, laundry basin, door. Not pictured: soakaway pit. Source: Water and Sanitation for the Urban Poor.

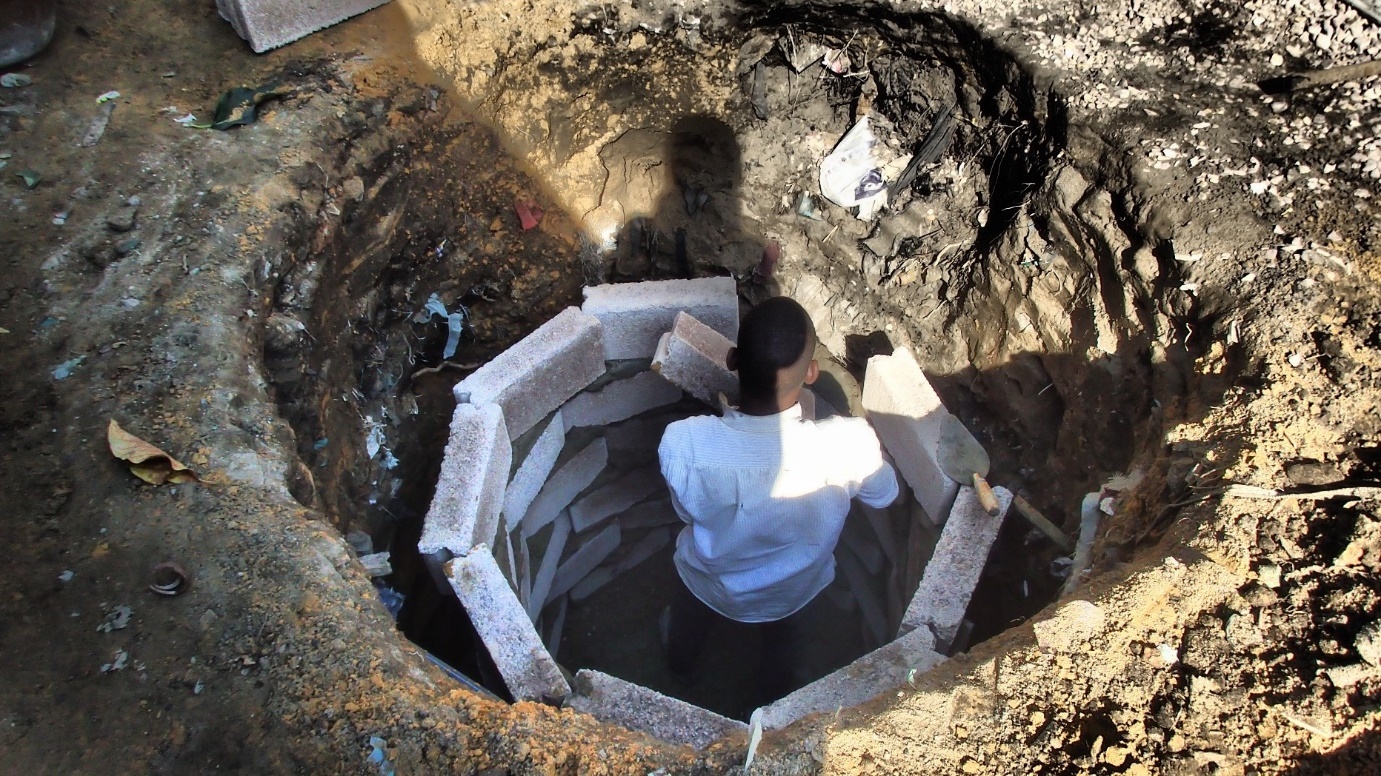

### Supplemental Figure 5b: Construction of a soakaway pit for discharge of liquid effluent from intervention latrines.

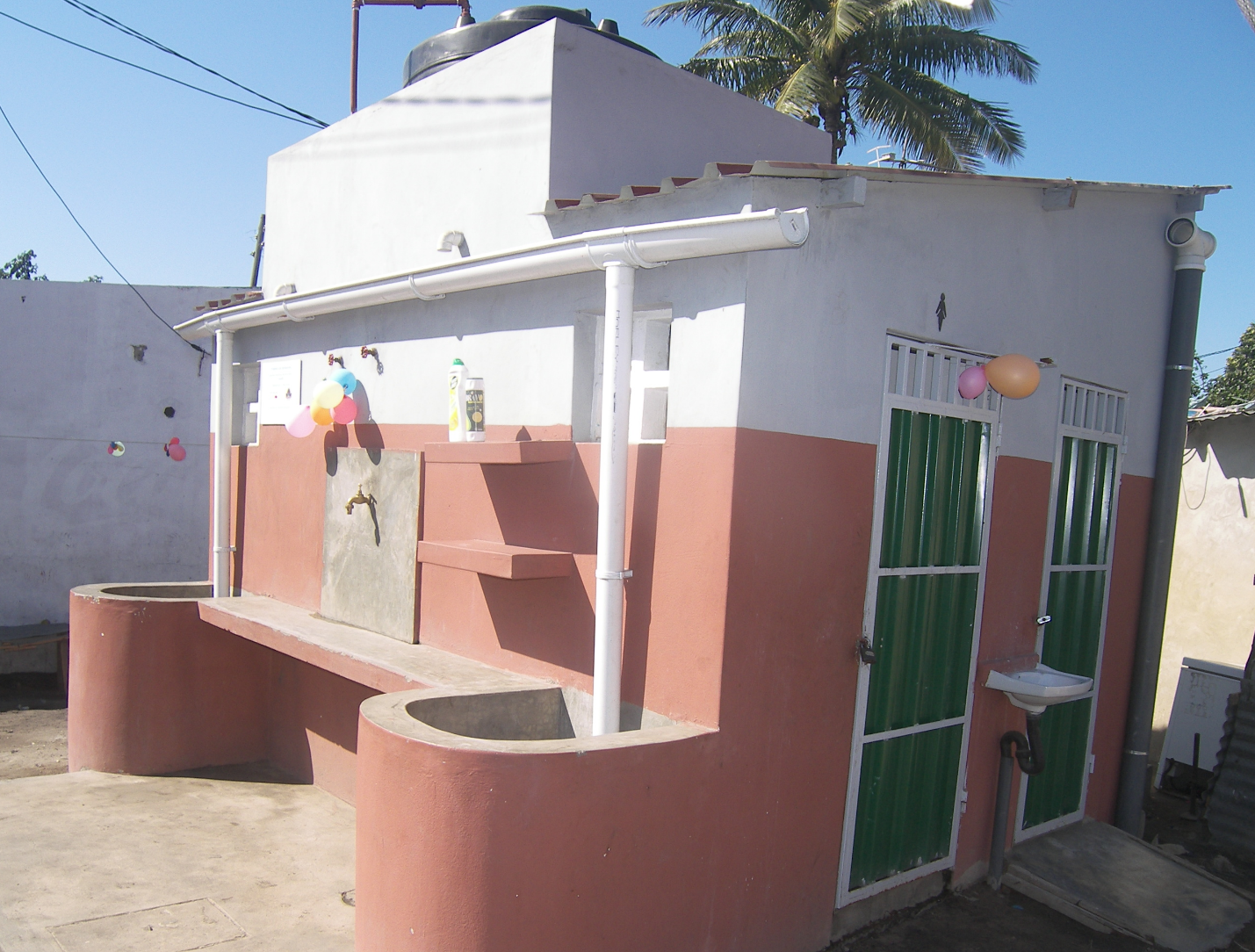

### Supplemental Figure 6a: Photo of communal sanitation block as constructed

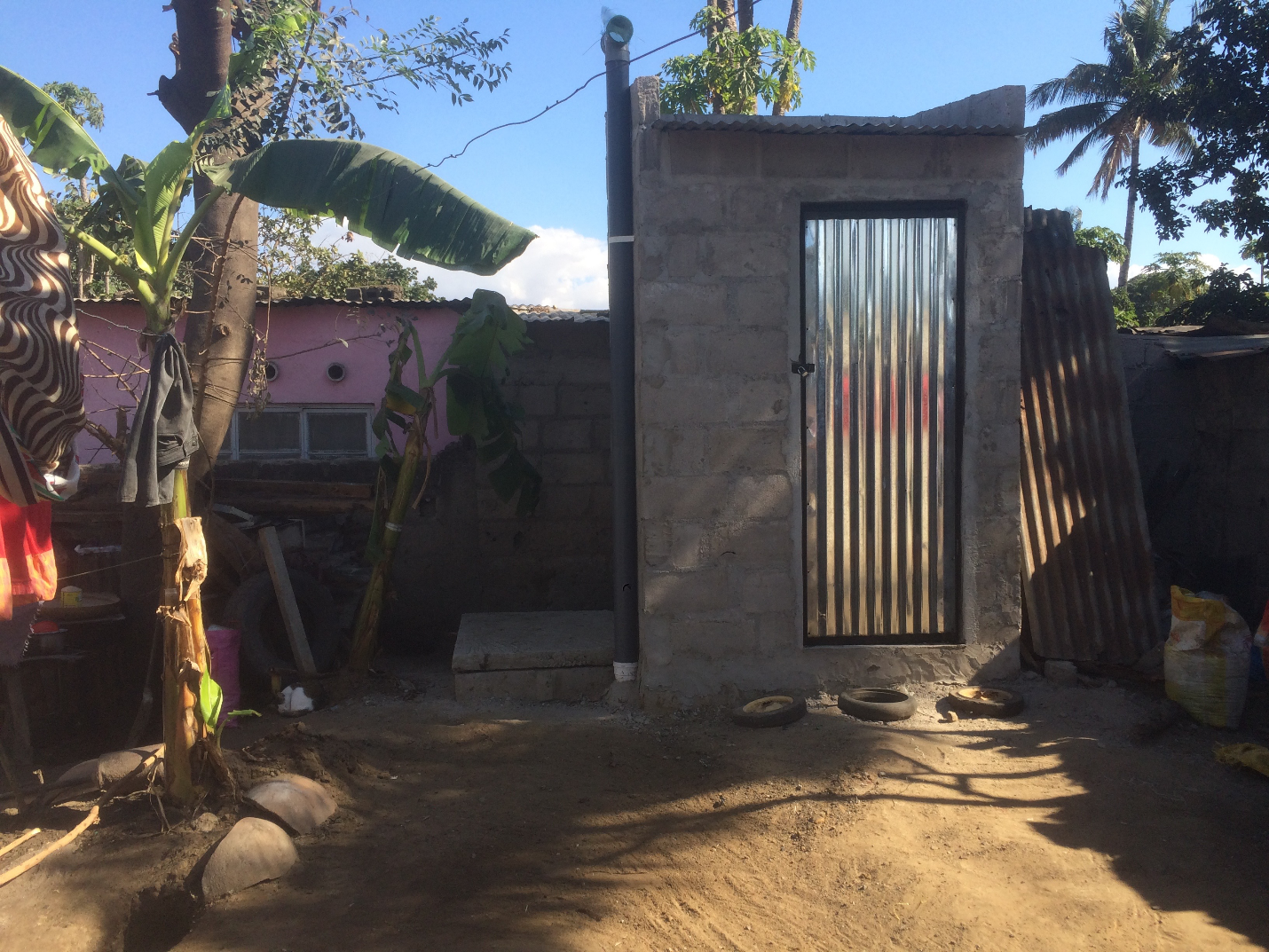

### Figure 6b: Photo of shared latrine as constructed

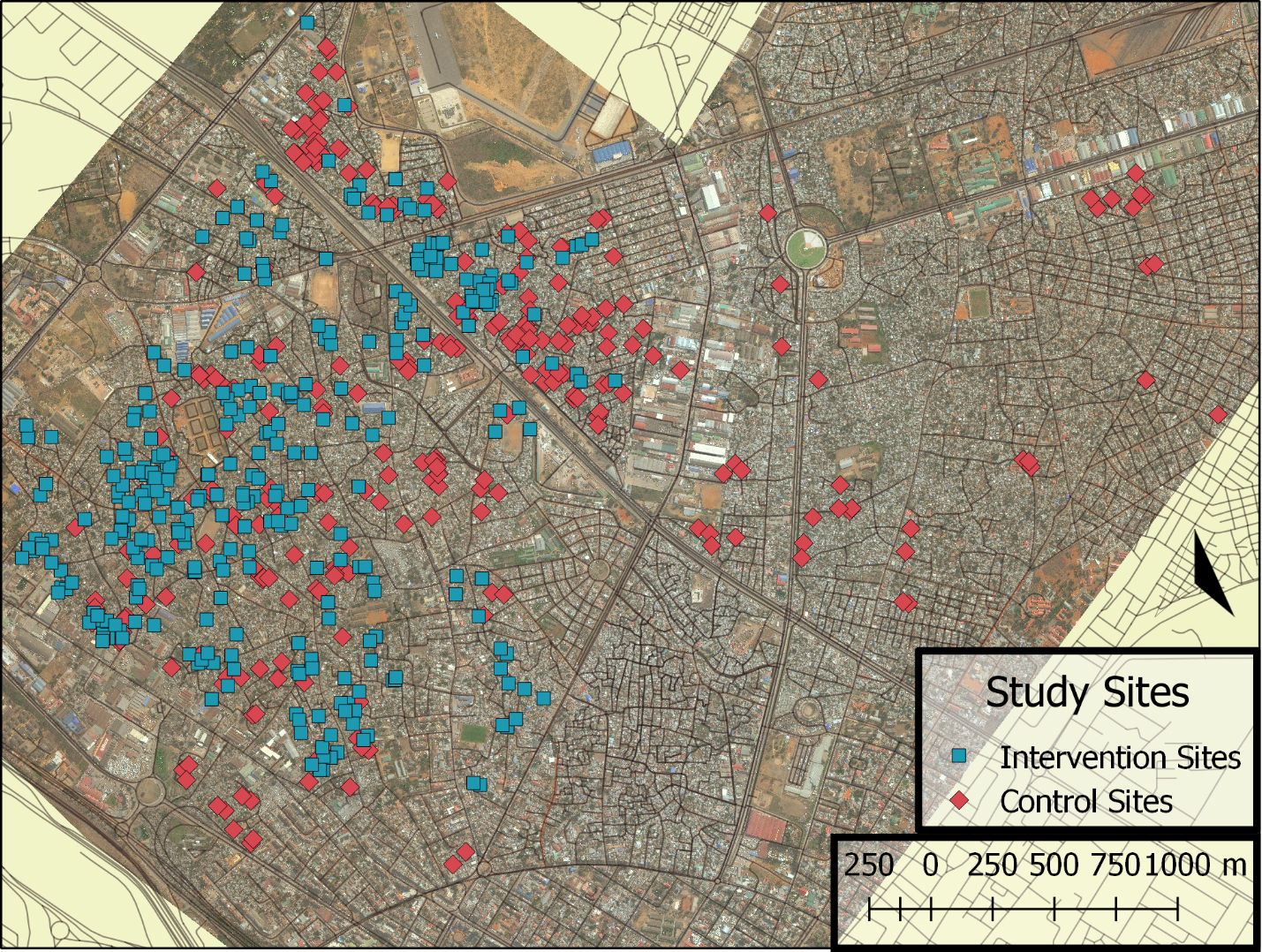

### Supplemental Figure 7: Map illustrating locations of intervention (n=208) and control sites (n=287) (compounds).

### Supplemental Methods: Consent procedures, survey administration, and specimen collection and analysis

Enumerators visited households with enrolled children at least twice at each point of follow-up. On the first visit of each phase, enumerators completed consent procedures, administered child-, household-, and compound-level surveys, and delivered stool sample collection supplies. The child’s mother was the target respondent for child and household surveys, though the father or another guardian was also eligible. For compound-level surveys, the head of the compound or his or her spouse was the preferred respondent. We sought written, informed consent from the parent or guardian of each eligible child prior to initial enrollment. We sought verbal assent from parents or guardians at each follow-up visit. Consent procedures, surveys, and all study-related verbal communication was performed in Portuguese or Changana as requested by the participant. Written materials were provided in Portuguese. Enumerators provided each caregiver with stool collection supplies, including disposable diapers, a plastic potty if the child was no longer wearing diapers, and a pre-labeled sterile sample bag. Enumerators returned the next day to collect the specimens. If a specimen was unavailable during the scheduled pickup, caregivers called the field team, using phone credit provided by the study, as soon as one was available or if fresh collection supplies were needed. If field enumerators were unable to collect a stool sample after multiple attempts, a registered nurse used an anatomically designed rectal swab (Copan Diagnostics Inc, Murrieta, CA, USA) to collect fecal material. Parents or guardians were required to complete a separate written consent procedure prior to collection of rectal swabs. Stool specimens and rectal swabs were stored in coolers with cold packs and delivered to the Medical Parasitology Laboratory at the Mozambican Ministry of Health (MISAU/INS) within six hours of collection. Technicians at INS prepared Kato-Katz slides for soil-transmitted helminth (STH) detection the day of receipt and read results within 30 minutes of preparation for hookworm and within 24 hours for other STH. In addition to STH analysis, laboratory technicians at INS also aliquoted stools into several sterile tubes and stored them, and any rectal swabs, at -80°C. If a child produced a liquid stool, lab technicians stored a piece of the saturated diaper material (“diaper samples”) at -80°C. Stool samples were shipped frozen on dry ice with temperature probes to the Georgia Institute of Technology in Atlanta, Georgia, USA where they were stored at -80°C until analysis.

We followed manufacturer instructions for the pretreatment, extraction, and analysis of stool samples by the Luminex Gastrointestinal Pathogen Panel (GPP), with additional elution steps added to the pretreatment protocol for rectal swabs and diaper samples. We eluted diaper samples in 2.5 mL of lysis buffer (ASL buffer, Qiagen, Hilden, Germany). We used a sterile 10-mL syringe to facilitate elution via agitation by taking in and expelling the buffer 5 times. We used 1 mL of the final eluate in the pretreatment. We agitated rectal swabs in 1 mL of lysis buffer for 1 minute and used the eluate in the pretreatment. Following pretreatment, we extracted DNA and RNA using the QIAcube HT platform and the QIAamp 96 Virus QIAcube HT Kit (Qiagen, Hilden, Germany). We added MS2, a non-pathogenic RNA virus, to each sample prior to nucleic acid extraction as an extraction and RT-PCR inhibition control. We included at least one sample process control (containing only lysis buffer and MS2) and negative extraction control (containing only lysis buffer) with each set of extractions. During the PCR step, we included at least one no-template control, containing molecular grade water and all PCR reagents, with each run. To assess elution and extraction of nucleic acid from swab and diaper samples, we measured the concentration of double-stranded DNA (dsDNA) present in a subset of extracts using the Qubit® High Sensitivity dsDNA kit (Invitrogen™, Carlsbad, CA, USA) and Qubit® 4 Fluorimeter (Invitrogen™, Carlsbad, CA, USA). The mean concentration of dsDNA recovered from rectal swabs was 26.3 ng/μL (SD 15.5, n=195, 25 swabs with measures above assay detection limit) and from diaper samples was 28.7 ng/μL (SD 16.9, n=61, 16 diapers with measures above assay detection limit). The concentration of dsDNA recovered from whole stool exceeded the assay detection limits in most cases. The mean concentration of dsDNA in the subset of stools with measurable results was 40.8 ng/μL (SD=16.5, n=33, 57 samples had concentrations above the assay detection limit). Following extraction, we stored all extracts at 4°C and analyzed them by GPP within 24 hours. For long-term storage, we archived samples at -80°C. We extracted and analyzed approximately 10% of samples in duplicate (biological replicates). If duplicate analyses yielded different results, we combined the results from all analyses such that the final result captured all positive detections for a given sample. If we could not detect a MS2 signal in a given sample, we either re-extracted or diluted the extract 1:10 in molecular grade water and re-assayed by GPP.

### Supplemental Table 16: Outcome and covariate descriptions, coding, and % missing.

|  | **Baseline, n=987** | **12-month, n=939** | **24-month, n=1001** |  |  |
| --- | --- | --- | --- | --- | --- |
|  | **% missing** | **% missing** | **% missing** | **Variable description** | **Data source** |
| **Outcome Data** |  |  |  |  |  |
| Enteric infection outcome data available | 24 | 14 | 8.0 | Binary; 0/1 | Based on collection of stool material and successful analysis by GPP |
| STH infection outcome data available | 30 | 37 | 46 | Binary; 0/1 | Based on collection of stool material and successful analysis by Kato-Katz |
| Caregiver-reported diarrhea, 7-day recall | 1.3 | 7.8 | 20 | Binary; 0/1 | Child Survey |
| **Covariate data** |  |  |  |  |  |
| Child sex, female | 2.3 | 1.3 | 7.0 | Binary; 0=male, 1=female | Child Survey |
| Respondent is child's mother | 2.5 | 7.6 | 20 | Binary; 0/1 | Child Survey |
| Caregiver completed primary school | 0.8 | 1.7 | 6.7 | Binary; 0/1 | Child Survey |
| Child breast feeds with or without complementary feeding | 1.3 | 7.7 | 20 | Binary; 0/1 | Child Survey |
| Child exclusively breastfeeds | 1.3 | 7.7 | 20 | Binary; 0/1 | Child Survey |
| Child wears a diaper | 1.4 | 7.6 | 20 | Binary; 0/1 | Child Survey |
| Child feces is disposed of in a latrine | 1.3 | 7.1 | 20 | Binary; 0/1 | Created from survey questions in Child Survey |
| Child age at sampling, days | 23 | 16 | 17 | Integer | Created from birthdate (Child Survey) and date of sampling |
| Child age at survey, days | 2.6 | 7.5 | 19 | Integer | Created from birthdate (Child Survey) and date of Survey |
| 30-day cumulative rainfall at sampling | 21 | 14 | 10 | Continuous | Created from sample date and data from data from the National Oceanic and Atmospheric Administration’s National Centers for Environmental Information (https://www.ncdc.noaa. gov/cdo-web/datatools/findstation) |
| 30-day cumulative rainfall at survey | 1.3 | 7.1 | 19 | Continuous | Created from survey date and data from data from the National Oceanic and Atmospheric Administration’s National Centers for Environmental Information (https://www.ncdc.noaa. gov/cdo-web/datatools/findstation) |
| Sample collection during rainy season | 21 | 14 | 10 | Binary; 0/1 | Created from sample date. Rainy season defined as November – April. |
| Survey collection during rainy season | 1.3 | 7.1 | 19 | Binary; 0/1 | Created from survey date. Rainy season defined as November – April. |
| Household crowding, >3 persons/room | 0.4 | 0.3 | 2.7 | Binary; 0/1 | Created from questions in Household Survey |
| Household floor is covered | 0.4 | 0.3 | 2.7 | Binary; 0/1 | Observation |
| Household walls made of concrete, bricks or similar | 0.4 | 0.3 | 2.7 | Binary; 0/1 | Observation |
| Household population | 0.3 | 0.3 | 1.6 | Integer | Household survey |
| Number of rooms in household | 0.4 | 0.3 | 2.3 | Integer | Created from questions in Household Survey |
| Wealth score, 0 (poorest) - 1 (wealthiest), unitless | 0.4 | 0.3 | 2.7 | Continuous | Created from questions in Household Survey using Simple Poverty Scorecard for Mozambique (http://www.simplepovertyscorecard.com/MOZ_2008_ENG.pdf). Questions referencing latrine removed from 12-month and 24-month score. All scores normalized by total number of points available. |
| Household uses tap in compound as primary drinking water source | 1.7 | 1.0 | 2.0 | Binary 0/1 | Created from drinking water source question in Household Survey |
| Latrine has drop-hole cover | 1.9 | 0.0 | 0.0 | Binary; 0/1 | Observation |
| Latrine has a ventpipe | 1.8 | 0.0 | 0.0 | Binary; 0/1 | Observation |
| Latrine has a ceramic, tile, or concrete pedestal or slab | 2.2 | 0.1 | 0.1 | Binary; 0/1 | Observation |
| Latrine has sturdy walls made of concrete, bricks, or similar. | 1.9 | 0.0 | 0.0 | Binary; 0/1 | Observation |
| Compound population | 0.0 | 0.0 | 0.0 | Integer | Compound Survey, enrollment checklists |
| Number of households in compound | 0.0 | 0.0 | 0.0 | Integer | Compound Survey, enrollment checklists |
| Number of latrines present in the compound | 0.1 | 0.0 | 0.0 | Integer | Compound Survey |
| Persons per latrine | 1.8 | 0.1 | 0.3 | Continuous | Created by dividing the compound population by the number of latrines/drop-holes |
| Households per latrine | 1.8 | 0.1 | 0.3 | Continuous | Created by dividing the number of households in the compound by the number of latrines in the compound |
| Number of water taps present in the compound | 1.1 | 0.0 | 0.0 | Integer | Compound Survey |
| Standing water visible around compound grounds | 1.9 | 0.3 | 0.0 | Binary; 0/1 | Observation |
| Standing or leaking wastewater visible around compound grounds | 1.9 | 0.3 | 0.0 | Binary; 0/1 | Observation |
| Faeces or used diapers observed around compound grounds or in solid waste | 1.9 | 0.3 | 0.0 | Binary; 0/1 | Observation |
| Compound floods when it rains | 0.0 | 0.0 | 0.0 | Binary; 0/1 | Compound Survey |
| Compound has electricity that normally functions | 0.0 | 0.0 | 0.0 | Binary; 0/1 | Compound Survey |
| Compound-level population density | 2.2 | 1.5 | 1.5 | Continuous, persons/m^2^ | Created by dividing the population of the compound by the measured area of the compound |
| Any animal present in the compound | 0.0 | 0.4 | 0.0 | Binary; 0/1 | Observation |
| Dog(s) present in the compound | 0.0 | 0.4 | 0.0 | Binary; 0/1 | Observation |
| Chicken(s) and/or duck(s) present in the compound | 0.0 | 0.4 | 0.0 | Binary; 0/1 | Observation |
| Cat(s) present in the compound | 0.0 | 0.4 | 0.0 | Binary; 0/1 | Observation |
| Any other animal(s) present in the compound | 0.0 | 0.4 | 0.0 | Binary; 0/1 | Observation |

Source files available in Supplemental Table 16 – source data 1 and Supplemental Table 16 – source code 1.

### Supplemental Table 17: Consort 2010 Checklist Extension for Cluster Trials.

| Section/Topic | Item No | Standard Checklist item | Extension for cluster designs | Page No * |
| --- | --- | --- | --- | --- |
| Title and abstract | | | |  |
|  | 1a | Identification as a randomized trial in the title | Identification as a cluster randomized trial in the title | N/A |
|  | 1b | Structured summary of trial design, methods, results, and conclusions (for specific guidance see CONSORT for abstracts)^[1],^^[2]^ | See table 2 | N/A based on journal’s abstract guideline. |
| Introduction | | | |  |
| Background and objectives | 2a | Scientific background and explanation of rationale | Rationale for using a cluster design | 4-5 |
|  | 2b | Specific objectives or hypotheses | Whether objectives pertain to the the cluster level, the individual participant level or both | 5 |
| Methods | | | |  |
| Trial design | 3a | Description of trial design (such as parallel, factorial) including allocation ratio | Definition of cluster and description of how the design features apply to the clusters | 28 |
|  | 3b | Important changes to methods after trial commencement (such as eligibility criteria), with reasons |  | **N/A** |
| Participants | 4a | Eligibility criteria for participants | Eligibility criteria for clusters | 30-31 |
|  | 4b | Settings and locations where the data were collected |  | **29-30** |
| Interventions | 5 | The interventions for each group with sufficient details to allow replication, including how and when they were actually administered | Whether interventions pertain to the cluster level, the individual participant level or both | 28-29 |
| Outcomes | 6a | Completely defined pre-specified primary and secondary outcome measures, including how and when they were assessed | Whether outcome measures pertain to the cluster level, the individual participant level or both | 33 |
|  | 6b | Any changes to trial outcomes after the trial commenced, with reasons |  | **33** |
| Sample size | 7a | How sample size was determined | Method of calculation, number of clusters(s) (and whether equal or unequal cluster sizes are assumed), cluster size, a coefficient of intracluster correlation (ICC or *k*), and an indication of its uncertainty | 33-34 |
|  | 7b | When applicable, explanation of any interim analyses and stopping guidelines |  | **N/A** |
| Randomisation: | | | |  |
| Sequence generation | 8a | Method used to generate the random allocation sequence |  | **N/A** |
|  | 8b | Type of randomisation; details of any restriction (such as blocking and block size) | Details of stratification or matching if used | N/A |
| Allocation concealment mechanism | 9 | Mechanism used to implement the random allocation sequence (such as sequentially numbered containers), describing any steps taken to conceal the sequence until interventions were assigned | Specification that allocation was based on clusters rather than individuals and whether allocation concealment (if any) was at the cluster level, the individual participant level or both | N/A |
| Implementation | 10 | Who generated the random allocation sequence, who enrolled participants, and who assigned participants to interventions | Replace by 10a, 10b and 10c |  |
|  | 10a |  | Who generated the random allocation sequence, who enrolled clusters, and who assigned clusters to interventions | 28-30 |
|  | 10b |  | Mechanism by which individual participants were included in clusters for the purposes of the trial (such as complete enumeration, random sampling) | 28-30 |
|  | 10c |  | From whom consent was sought (representatives of the cluster, or individual cluster members, or both), and whether consent was sought before or after randomisation | 30-31 |
| Blinding | 11a | If done, who was blinded after assignment to interventions (for example, participants, care providers, those assessing outcomes) and how |  | **30** |
|  | 11b | If relevant, description of the similarity of interventions |  | **28-29** |
| Statistical methods | 12a | Statistical methods used to compare groups for primary and secondary outcomes | How clustering was taken into account | 34-35 |
|  | 12b | Methods for additional analyses, such as subgroup analyses and adjusted analyses |  | **33-35** |
| Results | | | |  |
| Participant flow (a diagram is strongly recommended) | 13a | For each group, the numbers of participants who were randomly assigned, received intended treatment, and were analysed for the primary outcome | For each group, the numbers of clusters that were randomly assigned, received intended treatment, and were analysed for the primary outcome | Figure 1 |
|  | 13b | For each group, losses and exclusions after randomisation, together with reasons | For each group, losses and exclusions for both clusters and individual cluster members | Figure 1 |
| Recruitment | 14a | Dates defining the periods of recruitment and follow-up |  | **6** |
|  | 14b | Why the trial ended or was stopped |  | **N/A** |
| Baseline data | 15 | A table showing baseline demographic and clinical characteristics for each group | Baseline characteristics for the individual and cluster levels as applicable for each group | Table 1 |
| Numbers analysed | 16 | For each group, number of participants (denominator) included in each analysis and whether the analysis was by original assigned groups | For each group, number of clusters included in each analysis | Figure 1, Tables 2 and 3 |
| Outcomes and estimation | 17a | For each primary and secondary outcome, results for each group, and the estimated effect size and its precision (such as 95% confidence interval) | Results at the individual or cluster level as applicable and a coefficient of intracluster correlation (ICC or k) for each primary outcome | Table 2 |
|  | 17b | For binary outcomes, presentation of both absolute and relative effect sizes is recommended |  | Table 2, relative only |
| Ancillary analyses | 18 | Results of any other analyses performed, including subgroup analyses and adjusted analyses, distinguishing pre-specified from exploratory |  | Table 3, Supplemental Tables 12-15 |
| Harms | 19 | All important harms or unintended effects in each group (for specific guidance see CONSORT for harms^[3]^) |  | **N/A** |
| Discussion | | | |  |
| Limitations | 20 | Trial limitations, addressing sources of potential bias, imprecision, and, if relevant, multiplicity of analyses |  | **23-27** |
| Generalisability | 21 | Generalisability (external validity, applicability) of the trial findings | Generalisability to clusters and/or individual participants (as relevant) | 27-28 |
| Interpretation | 22 | Interpretation consistent with results, balancing benefits and harms, and considering other relevant evidence |  | **14-15, 27-28** |
| Other information | | |  |  |
| Registration | 23 | Registration number and name of trial registry |  | **35** |
| Protocol | 24 | Where the full trial protocol can be accessed, if available |  | **28** |
| Funding | 25 | Sources of funding and other support (such as supply of drugs), role of funders |  | **36** |
